# One night of closed-loop auditory stimulation during sleep enhances memory consolidation in patients with Alzheimer’s Disease

**DOI:** 10.64898/2026.09.14.26360893

**Authors:** Eduardo López-Larraz, David Oriol, Galit Fierro, Esperanza Jubera-García, Jens G. Klinzing, María Sierra-Torralba, Eduardo Horna-Prat, Carlos Escolano, Javier Sutil-Jiménez, Francisca Rojas, Luis Montesano, Cristina Moreno Loscertales, Gerard Piñol, Alvaro Pascual-Leone, Eugenia Marta Moreno, Elena Muñoz-Farjas, Javier Minguez

**Affiliations:** Bitbrain, Zaragoza, Spain; Department of Psychology, University of Cambridge, Cambridge, UK; 3Department of Computer Science and Systems Engineering (DIIS), University of Zaragoza, Zaragoza, Spain; Aragón Institute for Engineering Research (I3A), Zaragoza, Spain; Aragón Health Research Institute (IIS), Zaragoza, Spain; Hospital Universitario Miguel Servet, Zaragoza, Spain; Hospital Royo Villanova, Zaragoza, Spain; Cognition and Behaviour Study Group, Institut de Recerca Biomèdica de Lleida – Fundació Dr. Pifarré, IRBLleida, Lleida, Spain; Unitat de Trastorns Cognitius, Hospital Universitari Santa Maria de Lleida, Universitat de Lleida, Lleida, Spain; Department of Experimental Medicine, Faculty of Medicine, Universitat de Lleida, Lleida, Spain; Hinda and Arthur Marcus Institute for Aging Research, Deanna and Sidney Wolk Center for Memory Health, Hebrew SeniorLife, Boston, MA, USA; Department of Neurology, Harvard Medical School, Boston, MA, USA; Hospital Clínico Universitario Lozano Blesa, Zaragoza, Spain

**Keywords:** Alzheimer’s disease, Sleep, Closed-loop auditory stimulation (CLAS), Slow oscillations, Sleep spindles, Mild cognitive impairment, Memory consolidation, Declarative memory, Electroencephalography, Neuromodulation, Randomized controlled trial

## Abstract

**INTRODUCTION:** Sleep-dependent memory consolidation is impaired early in Alzheimer’s disease (AD). Closed-loop auditory stimulation (CLAS) of sleep slow oscillations enhances memory consolidation in healthy individuals, but its efficacy in AD remains unknown.

**METHODS:** We conducted a randomized, double-blind, placebo-controlled crossover trial in 37 patients with biomarker-confirmed early symptomatic AD and an amnestic clinical phenotype. We assessed the effect of one night of CLAS, compared with one night of sham stimulation, on declarative memory retention, sleep electrophysiology, and sleep architecture.

**RESULTS:** CLAS significantly improved overnight word-pair consolidation (Cohen’s d_z_ = 0.42, *p* = 0.04), enhanced stimulation-evoked slow-wave and fast spindle activity, and preserved sleep architecture and subjective sleep quality.

**DISCUSSION:** This study provides the first demonstration that CLAS modulates sleep electrophysiology and improves overnight memory consolidation in patients with biomarker-confirmed AD. Further studies need to address the sustainability of such effects with multi-night interventions for a clinically meaningful intervention.

## 1 Introduction

Alzheimer’s disease (AD) is the leading cause of dementia worldwide and represents a major and growing public health challenge as populations age [1]. Progressive impairment of declarative memory is among its earliest and most disabling cognitive features, reflecting early dysfunction of hippocampal and medial temporal lobe networks [2–4]. Sleep plays a fundamental role in the consolidation of newly acquired declarative memories through the coordinated interaction of slow oscillations (SOs), thalamocortical spindles, and hippocampal sharp-wave ripples during non-rapid eye movement (NREM) sleep [5–7]. Sleep disturbances are highly prevalent across the AD continuum [8], and are increasingly recognized as both a consequence and a contributor to disease progression through their bidirectional relationship with amyloid and tau pathology [9,10]. In particular, slow-wave sleep is markedly reduced from the earliest stages of the disease [11,12], potentially compromising SO-dependent memory consolidation [13,14]. Together, these observations provide a strong mechanistic rationale for therapeutic strategies aimed at restoring physiological slow oscillatory activity during sleep to preserve memory function and mitigate cognitive decline in individuals with AD.

Closed-loop auditory stimulation (CLAS) is a non-invasive approach designed to enhance endogenous SOs during sleep by delivering brief auditory stimuli phase-locked to the up-state of ongoing oscillations. In cognitively-unimpaired young and older adults, CLAS reliably enhances slow-wave activity and sleep spindle power, and is associated with improved declarative memory consolidation [15–19]. In individuals with cognitive impairment, a single night of stimulation has been shown to increase slow-wave activity with concomitant next-morning improvements in memory performance, whereas repeated stimulation across multiple nights has been associated with favorable changes in amyloid-related biomarkers [20,21]. In patients with AD, a pilot study has demonstrated that CLAS enhances slow-wave activity, despite substantial inter-individual variability in stimulation responsiveness [22,23]. Collectively, these findings indicate that the neural mechanisms underlying slow oscillations remain sufficiently preserved to be therapeutically engaged across the AD continuum.

Despite these encouraging findings, whether CLAS produces reliable improvements in memory performance in patients with AD remains to be determined. This question is particularly relevant because the cortical, thalamocortical, and hippocampal–neocortical networks supporting slow-wave generation, spindle activity, and sleep-dependent memory consolidation are affected even at the prodromal stages of the disease [4,10,13]. This study reports the results of a randomized, double-blind, placebo-controlled crossover trial evaluating the effects of a single night of CLAS in patients with biomarker-confirmed early symptomatic AD and an amnestic clinical phenotype. The primary objective was to determine whether CLAS improved declarative memory consolidation relative to sham stimulation. We additionally examined the acute electrophysiological effects of stimulation on SOs and spindle activity, and explored the relationship between stimulation-induced neural changes and memory performance.

## 2 Methods

### 2.1 Experiment design

This study was a randomized, double-blind, placebo-controlled crossover trial designed to evaluate the effects of CLAS during sleep in patients with biomarker-confirmed AD pathology and an amnestic clinical phenotype. The study targeted an early symptomatic AD population, with an amnestic mild cognitive impairment (MCI) phenotype as the primary clinical characterization at recruitment. Recruitment, neurological pre-screening, and eligibility assessments were conducted at the Neurology Departments of Hospital Universitario Miguel Servet, Hospital Clínico Universitario Lozano Blesa and Hospital Royo Villanova (Zaragoza, Spain). Experimental sessions were performed at the Bitbrain sleep laboratories (Zaragoza, Spain).

Patients were assigned to one of two condition orders (stimulation-first or sham-first) using a minimization algorithm [24] designed to balance allocation with respect to age, sex, and delayed recall performance on the Free and Cued Selective Reminding Test (FCSRT). Because recruitment and experimental sessions occurred concurrently, allocation was performed sequentially throughout enrolment. For each incoming pair of patients, the minimization algorithm compared both possible condition-order assignments and selected the one yielding the smaller between-group imbalance across the balancing variables. When both assignments produced equal imbalance, one was assigned at random. Condition assignment was stored in an encrypted file and administered automatically by the stimulation software, which delivered stimulation or sham according to each patient’s assigned order without revealing condition identity. Both patients and experimenters remained blind to condition allocation throughout the study.

The study was approved by the local Ethics Committee (Comité de Ética de la Investigación de la Comunidad Autónoma de Aragón; C.I. PI24/517), registered at ClinicalTrials.gov (NCT07402590), and conducted in accordance with the Declaration of Helsinki. All patients provided written informed consent prior to enrolment.

### 2.2 Patients

Eligible patients were adults meeting all of the following inclusion criteria: (1) age between 50 and 75 years; (2) native Spanish speaker; (3) normal or corrected-to-normal color vision; (4) diagnosis of Alzheimer’s disease, with an amnestic MCI phenotype according to the current clinical criteria of the National Institute on Aging–Alzheimer’s Association [25] and the International Working Group [26], as well as the clinical protocols of the participating hospitals; and (5) sufficient hearing without the use of hearing aids, verified by a hearing threshold below 50 dB during an audiometric assessment.

Exclusion criteria were: (1) unstable or severe chronic systemic disease, including advanced cardiovascular disease, advanced chronic kidney disease, severe pulmonary disease, metastatic or end-stage cancer, severe hematological disorders, or active uncontrolled infectious disease; (2) evidence of extensive microangiopathic cerebro-vascular disease, prior large vessel brain infarcts, or other neuroimaging findings potentially accounting for cognitive impairment; (3) alcohol or psychotropic substance abuse; (4) diagnosis of depression or any severe psychiatric disorder within the five years preceding evaluation; (5) changes in benzodiazepine or antidepressant treatment within the six months prior to initial evaluation; (6) current treatment with neuroleptics; (7) epilepsy requiring active treatment within the previous five years or any neurological comorbidity potentially associated with cognitive impairment; (8) illiteracy; and (9) moderate-to-severe sleep apnea requiring continuous positive airway pressure (CPAP) therapy during experimental nights.

Sample size was determined a priori using GPower 3.1.9.7 [27] for the main primary endpoint, i.e., the change in cognitive performance overnight in a declarative memory word-pair association task. Based on published meta-analyses of auditory stimulation effects on declarative memory [16], and given the limited available evidence in elderly and MCI populations, a conservative effect size of d = 0.5 was assumed. A minimum sample of n = 34 was estimated to achieve 80% power at α = 0.05 (two-tailed paired t-test, within-subjects design).

### 2.3 Experimental procedure and timeline

Following screening, each participant completed five laboratory visits over a 29-day period: one calibration night (Night 0), two experimental nights (Night 1 and Night 2), and two daytime long-term follow-up visits (LT1 and LT2) (Figure 1). Experimental nights were separated by a 14-day washout period and administered in randomized counterbalanced order. The sleep laboratory accommodated two patients simultaneously, in separate rooms, supervised overnight by a single experimenter.

**Figure 1.**
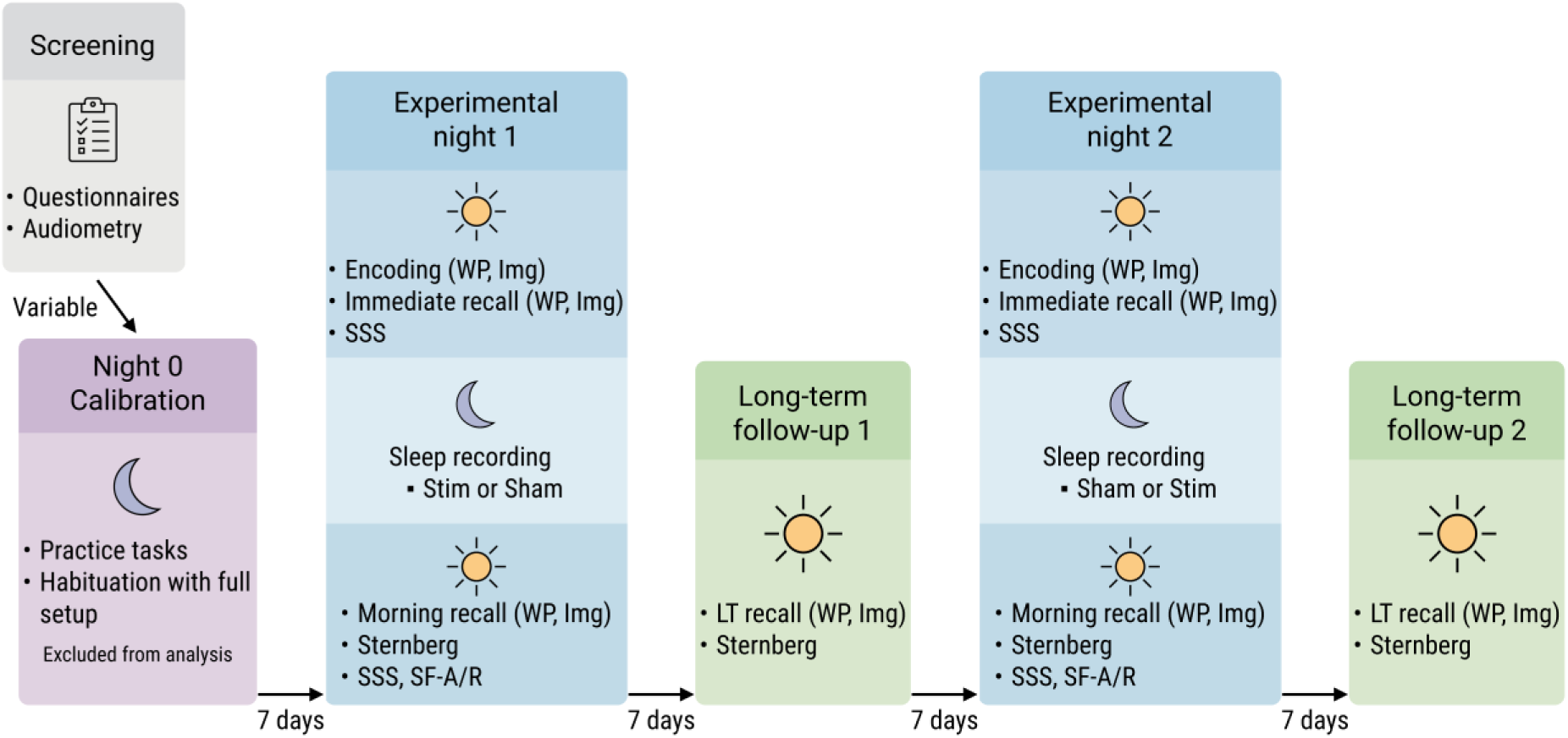
Experimental design and timeline. Schematic overview of the study protocol. Following screening, each participant completed five laboratory visits over 29 days. The calibration night (Night 0) served for habituation and stimulation parameter personalization and was excluded from analyses. Experimental nights included encoding and immediate recall before sleep, a sleep recording under auditory stimulation or sham conditions (counterbalanced), and morning cognitive assessment upon waking. Long-term follow-up visits comprised delayed recall of all memory tasks. Abbreviations: Img, visuospatial image recognition task; LT, long-term; SF-A/R, Schlaffragebogen A revidierte Fassung (subjective sleep questionnaire); SSS, Stanford Sleepiness Scale; WP, word-pair association task.

#### General overnight procedure

All overnight sessions followed the same structure. Patients arrived at the sleep laboratory at approximately 8:30 PM and completed intake questionnaires. The experimenter fitted the recording devices approximately one hour before sleep onset, and after that, patients performed the encoding and immediate retrieval portions of the declarative memory tasks. After that, the overnight recording started.

During the night, patients slept for approximately 7–8 hours while EEG and polysomnography (PSG) signals were recorded. The experimenter remained awake in an adjacent room throughout the night and supervised the recordings in real time. Infrared cameras installed in the rooms enabled continuous patient monitoring for safety purposes.

In the morning, patients completed post-sleep questionnaires, followed by the retrieval portions of the declarative memory tasks and assessment of working memory using the Sternberg task.

#### Screening and calibration night (Night 0)

Prior to the first laboratory night, patients attended a screening visit during which neurological eligibility was confirmed. Cognitive status was assessed using the Montreal Cognitive Assessment (MoCA) and the Free and Cued Selective Reminding Test (FCSRT). Patients also completed the Pittsburgh Sleep Quality Index (PSQI), as well as medical and demographic questionnaires. Hearing threshold was determined by audiometry and later used to define the initial auditory stimulation intensity.

The calibration night served both to habituate patients to the setup and protocol and to personalize stimulation parameters. During this night, patients performed abbreviated practice versions of the word-pair association and visuospatial image recognition tasks using a reduced number of stimuli. Auditory stimulation was delivered to assess individual electrophysiological responses and optimize stimulation settings, including slow oscillation detection thresholds and stimulation volume. Offline analysis of calibration-night recordings was subsequently used to finalize individualized stimulation parameters for the experimental nights (see Section 2.5).

#### Experimental nights (Night 1 and Night 2)

The first experimental night took place seven days after the calibration night, and the second experimental night was conducted after a 14-day washout period. Both nights followed identical procedures and differed only in the stimulation condition assignment.

During the stimulation night, CLAS was automatically delivered throughout stable NREM sleep following online slow oscillation detection. During the sham night, the full detection pipeline operated identically, but no auditory stimulation was delivered.

#### Long-term follow-up visits (LT1 and LT2)

Seven days after each experimental night, patients returned to the laboratory for a daytime follow-up session lasting approximately one hour. During these visits, patients completed delayed retrieval sessions for both declarative memory tasks, followed by the Sternberg working memory task.

### 2.4 Sleep recording system and auditory stimulation setup

Sleep was recorded using two concurrent systems. The primary recording device was the Ikon Sleep headband (Bitbrain, Spain), a wearable EEG system equipped with pre-gelled electrodes (Ambu, Denmark) positioned at AF7 and AF8, with ground and reference electrodes placed on the left and right mastoids, respectively. Signals were sampled at 256 Hz. The system additionally integrated photoplethysmography (PPG) for heart rate monitoring and a tri-axial accelerometer for head movement detection. Data were transmitted to a dedicated laptop via Bluetooth Low Energy. A custom software application running on the laptop acquired the EEG data stream for real-time CLAS.

Polysomnography was simultaneously recorded using the Micromed SD Plus system (Micromed, Italy), which captures electroencephalography, electrooculography, electromyography, electrocardiography, and respiratory activity via nasal airflow and thoracic respiratory belts. The system was connected to the same laptop via USB, and data were collected using dedicated acquisition software. PSG data were acquired for validation and future analyses but were not analyzed in the present study.

Auditory stimulation was delivered through two external speakers connected to the laptop via an audio jack cable, positioned on the headboard of the bed, approximately 50 cm apart and 20 cm above mattress level. Stimuli consisted of 50 ms bursts of pink noise, with volume automatically adjusted throughout the night to optimize the induction of slow oscillations while minimizing the risk of arousal (see Section 2.5).

### 2.5 Online detection and stimulation delivery

The software implemented three concurrent real-time processing loops on the EEG signal. The first loop performed continuous sleep stage classification using a deep neural network [28], trained on a sleep dataset previously acquired with the same device in a general population cohort [29]. The original five-class architecture was adapted into a binary classifier distinguishing stable deep sleep (N2–N3) from all other stages.

The second loop, active only during stable deep sleep, performed slow oscillation detection and phase-targeted stimulation delivery. The EEG was band-pass filtered between 0.2 and 5 Hz. Slow oscillations were detected by identifying zero-crossings from positive to negative and back to positive, with the intervening negative peak evaluated against an individualized amplitude threshold. Additional duration and amplitude checks were used to reject non-physiological events. For the calibration night, the threshold was set to a default of −60 µV, and subsequent offline analysis of this night was used to adjust this threshold toward zero until a minimum rate of 3 slow oscillations per minute was achieved. This adjusted threshold was then fixed for both experimental nights. Upon slow wave detection, a phase-locked loop (PLL) [30] tracked the instantaneous oscillation phase and issued a stimulation command when the estimated phase reached the slow wave up-state.

Because sleep staging operates on 30-second sliding windows with slow dynamics, a third loop implemented an arousal detector to respond rapidly to sudden changes in brain activity. This detector monitored short-term spectral changes in the EEG, particularly increases in beta power (20–30 Hz) relative to each individual’s baseline activity, which are indicative of micro-arousals or awakenings. When such an increase was detected, stimulation was immediately paused for a fixed period of 15 seconds to avoid further sleep disruption.

Stimulation was structured in alternating 6-second ON and OFF windows throughout stable deep sleep [19]. Detections during ON windows were stimulated (Stim ON condition); whereas detections during OFF windows were withheld (Stim OFF condition). On the sham night, the algorithm ran identically and detections were stored for analysis, but no stimulation was delivered (Sham night condition).

Stimulation volume was adapted throughout the night, remaining within ±8 dB relative to the initial value. The volume was increased by 2 dB when fewer than 2 of the last 10 actual stimulations were followed by a subsequent slow oscillation, and decreased by 4 dB when stimulation caused an arousal. The initial volume on the calibration night was set 8 dB above each patient’s hearing threshold from the screening session (with a range of 28–57 dB, corresponding to an overall range of 20–65 dB). Before the first experimental night, the optimal stimulation volume was selected based on offline analysis of calibration-night responses.

### 2.6 Sleep outcomes

#### 2.6.1 EEG signal processing and electrophysiological outcomes

All electrophysiological analyses were computed on the average of the two EEG channels recorded by the headband (AF7 and AF8), and all results are reported for this average signal. Prior to analysis, epochs were subjected to a multi-criterion artifact rejection procedure applied within a window of −2.5 to +3 seconds around each detected slow oscillation. Epochs were rejected if any of the following criteria were met: (1) peak amplitude exceeding 300 µV; (2) standard deviation of the low-frequency filtered signal (0.1–4 Hz) exceeding a data-driven threshold computed as the 75th percentile plus 1.5 times the interquartile range (Tukey fence) across all in-bounds epochs; or (3) standard deviation of the high-frequency filtered signal (40– 100 Hz) exceeding the same threshold, used as a proxy for muscular contamination. All event-locked analyses were time-locked to the slow oscillations detected online during the night, using an epoch window of −3 to +5 seconds relative to detection.

#### Slow oscillation morphology

For slow oscillation analyses, the EEG was bandpass filtered between 0.2 and 2 Hz (second-order zero-phase Butterworth filter). Grand average waveforms were computed across all artifact-free detected events per condition (Stim ON, Stim OFF, Sham night). Four amplitude descriptors were extracted from each detected event: the negative peak preceding stimulation (N1, searched in the [−1, 0] s window), the subsequent positive peak (P1, [0, 1] s), the stimulation-induced negative peak (N2, from P1 latency to 1.5 s), and the following positive peak (P2, from N2 latency to 2.0 s), identified as the local minima and maxima of the filtered signal within these predefined time windows. Peak-to-peak amplitudes (P1–N1, P1–N2, P2– N2) were also computed.

#### Time-frequency analysis

For time-frequency analyses, the EEG was bandpass filtered between 0.1 and 30 Hz and notch filtered at 48–52 Hz (second-order zero-phase Butterworth filters). Event-related spectral perturbation (ERSP) was computed using Morlet wavelets implemented in FieldTrip [31], with frequencies ranging from 0.5 to 20 Hz in 0.25 Hz steps and a time resolution of 50 ms. The number of wavelet cycles increased linearly from 3 cycles at 0.5 Hz to 10 cycles at 20 Hz, providing a frequency-dependent trade-off between temporal and spectral resolution. A baseline correction was applied using the mean power in the [−2.5, −2] second pre-detection window, and power was expressed in decibels (dB) relative to baseline. Band power was quantified by averaging baseline-corrected power within the [0, 2] second post-detection window across six frequency bands: slow oscillations (0.5–1 Hz), delta (1–4 Hz), theta (4–8 Hz), alpha (8–12 Hz), spindle (12–16 Hz), and beta (15–30 Hz).

#### Spindle activity

Fast and slow spindle activity was analyzed by bandpass filtering the EEG in the 12–16 Hz and 9–12 Hz ranges respectively (second-order zero-phase Butterworth filters), followed by amplitude envelope extraction using a 200 ms smoothing window. A detection threshold was defined as the 75th percentile of envelope power across all artifact-free samples of the recording. Epochs exceeding this threshold were classified as spindle events. Spindle co-occurrence with slow oscillations was assessed by identifying spindle events occurring within a ±250 ms window around the positive peak (P1) of each slow oscillation detected. A baseline correction was applied to spindle envelopes using the mean amplitude in the [−2.5, −2] second pre-detection window, consistent with the ERSP baseline.

#### 2.6.2 Sleep macrostructure

Sleep staging was performed automatically using the deep neural network described in Section 2.5, here applied in its original five-class configuration to produce standard hypnograms (Wake, N1, N2, N3, REM) [28], following American Academy of Sleep Medicine (AASM) guidelines [32]. This model achieves an agreement of over 85% with expert manual scoring [29], comparable to typical inter-rater agreement between human scorers [33]. Standard macrostructure metrics were extracted from the hypnograms, including total sleep time, sleep period time, sleep efficiency, wake after sleep onset, sleep onset latency, number of awakenings, and time and percentage spent in each sleep stage. Slow oscillation counts per condition (Stim ON, Stim OFF, Sham night) and coupled slow oscillation rates were also analyzed.

### 2.7 Cognitive outcomes

#### 2.7.1 Word-pair association task

Verbal episodic memory was assessed using a paired-associate word learning task. Each experimental night used a unique list of 30 semantically related word pairs to minimize interference across sessions. A shorter list of 15 word pairs was used on the calibration night for practice purposes.

During the evening learning phase, word pairs were presented sequentially on screen, with a 20-second rest period after every ten pairs. Immediately after learning, participants completed an immediate recall session in which only the left word of each pair was presented as a retrieval cue for 4 seconds, with a 1-second interstimulus interval, and participants had to verbally recall the associated word. A morning recall session was administered approximately one hour after waking, following the same procedure. A long-term recall session was conducted seven days after each experimental night, in which participants were again cued with the left words and asked to recall the associated words from that session.

Verbal responses were audio-recorded and scored offline by a researcher blinded to the condition of the experimental night. Each correctly recalled word pair was awarded one point. Memory retention was quantified as the number of correctly recalled pairs at morning or long-term recall minus the number recalled at immediate recall.

#### 2.7.2 Visuospatial image recognition task

Visuospatial episodic memory was assessed using an image recognition task. During the evening learning phase, 38 images of everyday objects or faces were presented sequentially for 2 seconds each, displayed in one of four screen quadrants. Participants were instructed to memorize both the image identity and its spatial location. Different image sets were used across experimental nights to minimize interference.

Memory retrieval was assessed at immediate recall, morning recall, and long-term recall. At each retrieval session, the 38 previously learned images were randomly intermixed with 38 novel images. Participants indicated whether they recognized each image and, for recognized images, identified the screen quadrant in which it had originally appeared. A shorter version using 6 learned and 6 novel images was administered during the calibration night for practice purposes. Each correctly classified image was awarded one point. A previously learned image received one point if both its identity and spatial location were correctly identified, whereas a novel image received one point if it was correctly rejected. Thus, the maximum score was 76 points. Memory retention was quantified as the number of correctly classified images at morning or long-term recall, minus performance at immediate recall.

#### 2.7.3 Sternberg working memory task

Working memory was assessed using an adapted version of the Sternberg task, administered on the morning following each experimental night and at long-term follow-up. This task served as a control to assess whether stimulation effects were specific to sleep-dependent declarative memory consolidation, given that working memory is not expected to benefit from slow-wave enhancement during sleep [16].

In each trial, a sequence of 5 or 6 digits was displayed for 3 seconds, followed by a 5-second retention interval. Three probe digits were then presented sequentially, and participants indicated whether each probe had been part of the preceding sequence. Each probe was displayed for 3 seconds, with a 3.5-second interstimulus interval between probes. Target and non-target probes were balanced at 50% within each trial, and serial position of target probes was counterbalanced. The task comprised 16 trials (8 five-digit and 8 six-digit sequences) with an approximate duration of 6 minutes. A shortened version of 4 trials was administered on the calibration night for practice. Accuracy, measured as the total number of correct responses, and reaction times were recorded.

#### 2.7.4 Subjective sleep measures

Subjective sleep quality was assessed after each overnight session using a Spanish translation of the Schlaffragebogen A, revidierte Fassung (SF-A/R) [34], a validated self-report questionnaire covering 10 indices of sleep experience including ease of falling asleep, sleep continuity, early awakening, general sleep characteristics, total sleep duration, sleep quality, feeling of recovery, psychological balance before sleep, psychological exhaustion before sleep, and psychosomatic symptoms during sleep. Subjective sleepiness was assessed before sleep onset and after waking using the Stanford Sleepiness Scale (SSS) [35], a 7-point scale ranging from full alertness to near sleep onset.

### 2.8 Statistical analysis

All statistical analyses were performed in MATLAB R2023a. Analyses of primary cognitive outcomes were pre-specified and considered confirmatory; secondary electrophysiological outcomes were pre-specified with directional hypotheses; all remaining analyses were exploratory and should be interpreted accordingly.

#### Primary cognitive outcomes

The effect of auditory stimulation on declarative episodic memory retention was assessed using linear mixed-effects models (LMM) fitted with MATLAB’s *fitlme* function, with the model formula *outcome ∼ condition + (1|patient)*, where *condition* was modeled as a two-level categorical fixed effect (stimulation night vs. sham night) and *patient* as a random intercept to account for the paired crossover structure. Normality of model residuals was verified using the Lilliefors test, and no violations were detected. Statistical inference for fixed effects was based on Wald *t*-tests using Satterthwaite’s approximation for denominator degrees of freedom. To control for multiple comparisons across the two primary outcome measures, Bonferroni correction was applied separately at each retention interval (overnight and long-term). For each model, results are reported as the fixed-effect estimate (*β*), its standard error (SE), the *t* statistic with denominator degrees of freedom in parentheses, and the corresponding *p*-value; corrected *p*-values are reported in the text, whereas figures display uncorrected values. Effect sizes are reported as Cohen’s d_z_, computed as the mean paired difference divided by the standard deviation of the paired differences. The same LMM framework was applied to assess night order effects and working memory outcomes, treated as exploratory analyses.

#### Electrophysiological outcomes

Stimulation effects on EEG metrics were assessed as pre-specified secondary outcomes across three conditions (Stim ON, Stim OFF, Sham night), with directional hypotheses predicting enhanced slow oscillation amplitude and spindle activity in the Stim ON condition relative to both control conditions. Prior to analysis, normality of pairwise differences was assessed using the Lilliefors test. Normally distributed metrics were analyzed using one-way repeated measures ANOVA; when Mauchly’s test indicated violation of sphericity (*p* < 0.05), the Greenhouse-Geisser correction was applied. Non-normally distributed metrics were analyzed using the Friedman test. For metrics yielding a significant omnibus effect, pairwise post-hoc comparisons were conducted using paired *t*-tests or Wilcoxon signed-rank tests as appropriate, with Bonferroni correction applied within each omnibus to control for the three pairwise comparisons. Uncorrected *p*-values are displayed in figures; corrected significance is reported in the text.

#### Time-frequency analysis

Pairwise differences in event-related spectral perturbation (ERSP) between conditions were assessed using cluster-based permutation testing [36] implemented in FieldTrip [31]. Dependent-samples *t*-statistics were computed at each time-frequency pixel and adjacent significant pixels were grouped. Cluster-level statistics were computed across 1000 random permutations, and the significance threshold was set at *p* < 0.001 (two-tailed).

#### Brain-behavior correlations

Associations between electrophysiological metrics and memory performance were assessed as exploratory analyses using Pearson or Spearman correlations depending on the normality of each variable, assessed via the Lilliefors test. No correction for multiple comparisons was applied. Findings are reported at nominal significance thresholds of *p* < 0.05 and *p* < 0.1 (trend level).

#### Sleep macrostructure, subjective measures, and environmental variables

Differences in sleep architecture metrics, subjective sleep quality and sleepiness indices, and environmental conditions (room temperature and humidity) between stimulation and sham nights were assessed using paired t-tests or Wilcoxon signed-rank tests depending on normality, assessed via the Lilliefors test. Given that these analyses were intended to assess whether stimulation produced systematic differences between conditions, uncorrected *p*-values are reported throughout.

## 3 Results

### 3.1 Patient characteristics

A total of 59 patients were assessed for eligibility. Following neurologist pre-screening, 39 participants were enrolled in the intervention phase. Of these, 37 were ultimately included in the analyses, after exclusion of 2 participants due to the late identification of exclusion criteria, specifically active treatment with neuroleptics (N = 1) and a recent change in antidepressant medication within the previous three months (N = 1). The recruitment, enrollment, and exclusion flow is summarized in Supplementary Figure 1.

Baseline demographic and clinical characteristics of the analyzed sample are summarized in Table 1. The two counterbalanced groups were comparable with respect to age, sex, body mass index, cognitive measures, sleep quality, and educational level. The mean MoCA score was 19.1 ± 5.5 (median, 20; range, 6–29), reflecting the variability in global cognitive performance commonly observed in early-stage AD populations. Most patients were receiving cholinesterase inhibitor treatment, predominantly donepezil (67.6%). Benzodiazepine use was present in only 5 patients, all of whom were in Group 1. Only one patient had a diagnosis of obstructive sleep apnea and did not use CPAP during the experimental nights. Individualized stimulation parameters derived from the calibration night were comparable between groups (SO detection threshold: −45.2 ± 10.3 µV; initial volume: 37.3 ± 6.2 dB).

**Table 1.**
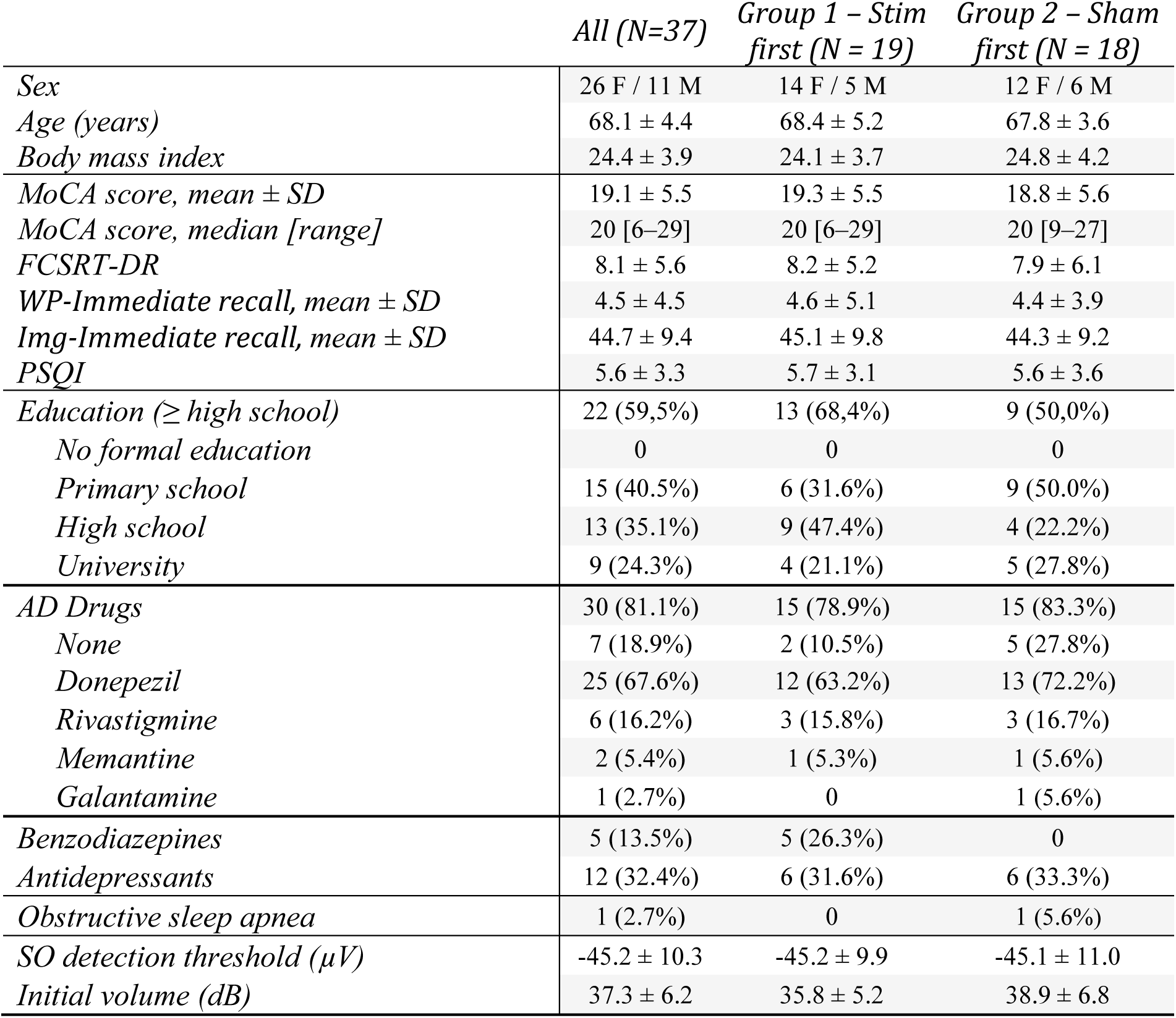
Baseline characteristics and individualized stimulation parameters. Values are mean ± SD or n (%) unless otherwise stated. Abbreviations: AD, Alzheimer’s disease; FCSRT-DR, Free and Cued Selective Reminding Test, delayed recall; Img-Immediate recall, average immediate recall of the two experimental nights in the image recognition task; MoCA, Montreal Cognitive Assessment; PSQI, Pittsburgh Sleep Quality Index; SO, slow oscillation; WP-Immediate recall, average immediate recall of the two experimental nights in the word-pair task.

|  | *All (N=37)* | *Group 1 – Stim*<br>*first (N = 19)* | *Group 2 – Sham*<br>*first (N = 18)* |
| --- | --- | --- | --- |
| *Sex* | 26 F / 11 M | 14 F / 5 M | 12 F / 6 M |
| *Age (years)* | 68.1 ± 4.4 | 68.4 ± 5.2 | 67.8 ± 3.6 |
| *Body mass index* | 24.4 ± 3.9 | 24.1 ± 3.7 | 24.8 ± 4.2 |
| *MoCA score, mean ± SD* | 19.1 ± 5.5 | 19.3 ± 5.5 | 18.8 ± 5.6 |
| *MoCA score, median [range]* | 20 [6–29] | 20 [6–29] | 20 [9–27] |
| *FCSRT-DR* | 8.1 ± 5.6 | 8.2 ± 5.2 | 7.9 ± 6.1 |
| *WP-Immediate recall, mean ± SD* | 4.5 ± 4.5 | 4.6 ± 5.1 | 4.4 ± 3.9 |
| *Img-Immediate recall, mean ± SD* | 44.7 ± 9.4 | 45.1 ± 9.8 | 44.3 ± 9.2 |
| *PSQI* | 5.6 ± 3.3 | 5.7 ± 3.1 | 5.6 ± 3.6 |
| *Education (≥ high school)* | 22 (59,5%) | 13 (68,4%) | 9 (50,0%) |
| *No formal education* | 0 | 0 | 0 |
| *Primary school* | 15 (40.5%) | 6 (31.6%) | 9 (50.0%) |
| *High school* | 13 (35.1%) | 9 (47.4%) | 4 (22.2%) |
| *University* | 9 (24.3%) | 4 (21.1%) | 5 (27.8%) |
| *AD Drugs* | 30 (81.1%) | 15 (78.9%) | 15 (83.3%) |
| *None* | 7 (18.9%) | 2 (10.5%) | 5 (27.8%) |
| *Donepezil* | 25 (67.6%) | 12 (63.2%) | 13 (72.2%) |
| *Rivastigmine* | 6 (16.2%) | 3 (15.8%) | 3 (16.7%) |
| *Memantine* | 2 (5.4%) | 1 (5.3%) | 1 (5.6%) |
| *Galantamine* | 1 (2.7%) | 0 | 1 (5.6%) |
| *Benzodiazepines* | 5 (13.5%) | 5 (26.3%) | 0 |
| *Antidepressants* | 12 (32.4%) | 6 (31.6%) | 6 (33.3%) |
| *Obstructive sleep apnea* | 1 (2.7%) | 0 | 1 (5.6%) |
| *SO detection threshold (μV)* | -45.2 ± 10.3 | -45.2 ± 9.9 | -45.1 ± 11.0 |
| *Initial volume (dB)* | 37.3 ± 6.2 | 35.8 ± 5.2 | 38.9 ± 6.8 |

### 3.2 Memory effects

#### 3.2.1 Stimulation effects on overnight memory consolidation

Five patients were excluded from word-pair task analyses due to inability to perform the task and/or non-compliance with task instructions. All word-pair analyses were therefore conducted on n = 32 patients. All 37 patients completed the visuospatial image recognition task and were included in the corresponding analyses.

Auditory stimulation during sleep significantly improved overnight retention in the word-pair task compared to sham (β = 0.88, SE = 0.37, t(62) = 2.39, *p* = 0.04, Bonferroni-corrected, Cohen’s d_z_= 0.42; Figure 2A). No significant effect was observed for the image recognition task (β = -0.65, SE = 1.04, t(72) = −0.62, *p* = 0.54, Cohen’s d_z_ = −0.10; Figure 2B).

**Figure 2.**
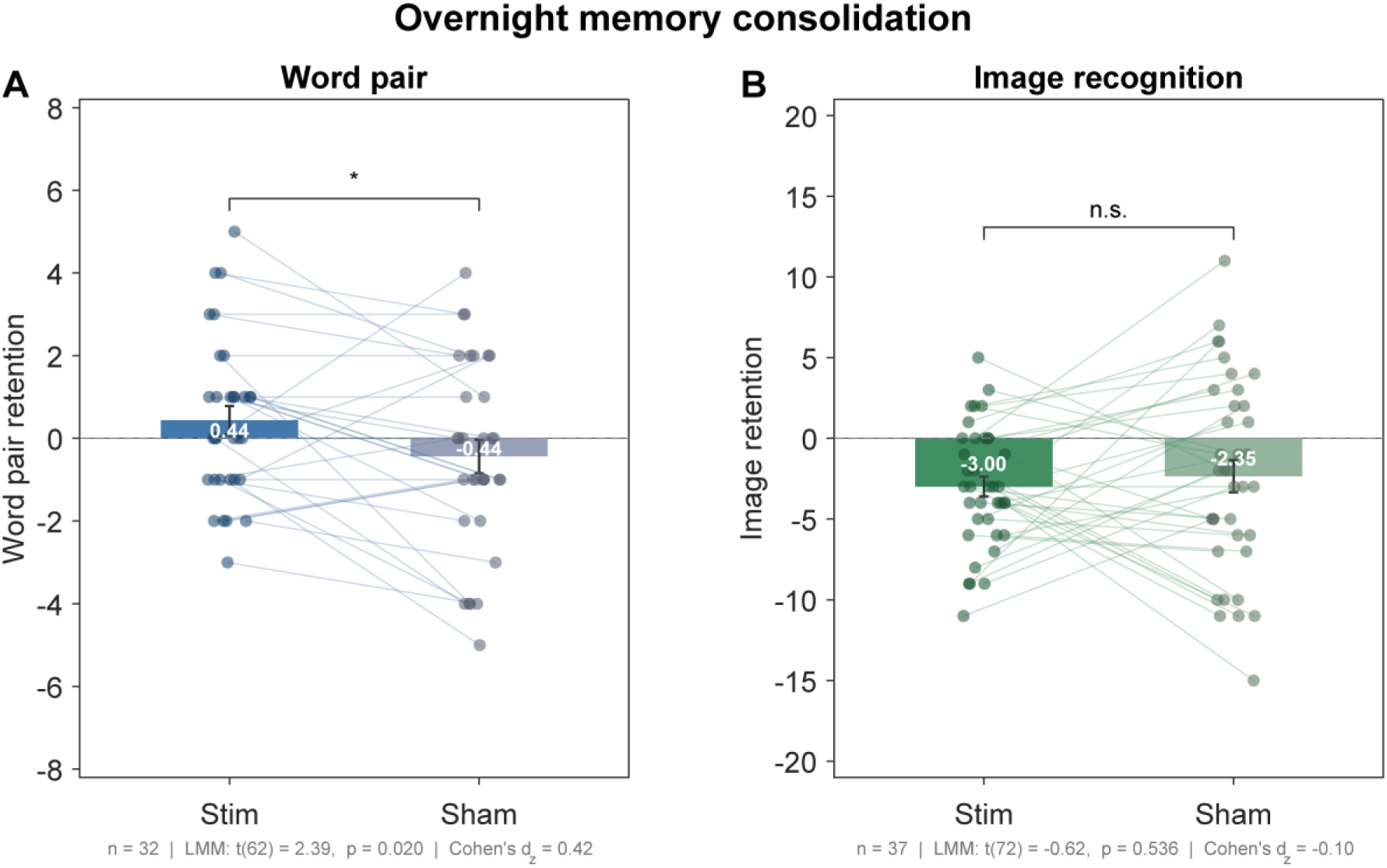
Effects of CLAS on overnight memory consolidation. (A) Overnight word-pair retention (morning − immediate recall) was enhanced in the stimulation vs the sham condition. (B) No significant difference was detected for the visuospatial image recognition task. n.s., not significant; *p < 0.05.

#### 3.2.2 Stimulation effects on long-term memory consolidation

Both long-term recall sessions were completed by 25 of the 32 patients included in the word-pair analysis. Seven individual sessions were lost due to a technical failure in the audio recording system, resulting in 26 stimulation and 31 sham sessions available for analysis.

No significant effect of stimulation was observed at the 7-day follow-up for either task. Word-pair consolidation did not differ between conditions (β = -0.01, SE = 0.57, t(55) = -0.01, *p* = 0.99, Cohen’s d_z_ = 0.00; Supplementary Figure 2A), nor did image recognition (β = -0.16, SE = 0.93, t(72) = −0.18, *p* = 0.86, Cohen’s d_z_ = −0.03; Supplementary Figure 2B).

#### 3.2.3 Night order effects on task performance

Night order effects were assessed by comparing performance between the first and second experimental nights, irrespective of condition assignment. No significant effect was observed for the word-pair task at either time point (morning: β = 0.12, SE = 0.40, t(62) = 0.31, *p* = 0.75, Cohen’s d_z_ = 0.05; 7-day follow-up: β = -0.04, SE = 0.57, t(55) = -0.06, *p* = 0.95, Cohen’s d_z_ = 0.05; Supplementary Figure 3A,C).

For the image recognition task, a numerically large but non-significant night order effect was observed at morning recall (β = 1.62, SE = 1.01, t(72) = 1.60, *p* = 0.11, Cohen’s d_z_ = 0.26), which reached significance at the 7-day follow-up (β = 2.59, SE = 0.82, t(72) = 3.16, *p* = 0.002, Cohen’s d_z_ = 0.51; Supplementary Figure 3B,D). Patients who completed the image recognition task on the second experimental night exhibited greater long-term forgetting, suggesting that night order may have represented a relevant source of variance for this task.

#### 3.2.4 Effects on working memory

No significant effects of auditory stimulation were observed for the Sternberg working memory task. Neither accuracy nor reaction time differed significantly between stimulation and sham conditions at either the overnight or 7-day follow-up assessments (all *p* > 0.05; Supplementary Figure 4).

Exploratory analyses of night order effects revealed a nominally significant increase in accuracy during the second night compared to the first (*p* = 0.03); however, this effect did not survive correction for multiple comparisons. No other night order effects were observed for either accuracy or reaction time (all *p* > 0.05; Supplementary Figure 4).

### 3.3 Sleep electrophysiology

The grand average SO waveform following auditory stimulation (Stim ON) reveals a prominent positive peak following the detected slow wave, succeeded by a second negative wave and a subsequent positive rebound (Figure 3A). Both Stim OFF and Sham Night were morphologically indistinguishable from each other and lacked this response. Statistical analyses confirmed significantly greater amplitudes of the stimulation-induced components (P1, N2, P2) relative to both Stim OFF and Sham Night conditions (Supplementary Figure 5). Fast spindle activity was also enhanced by auditory stimulation (Figure 3B), revealed as a significantly larger co-occurrence of spindles in Stim ON condition, as well as a significantly larger signal envelope amplitude during the 2 s period following stimulation (Supplementary Figure 5). No comparable effects were observed for slow spindles.

**Figure 3.**
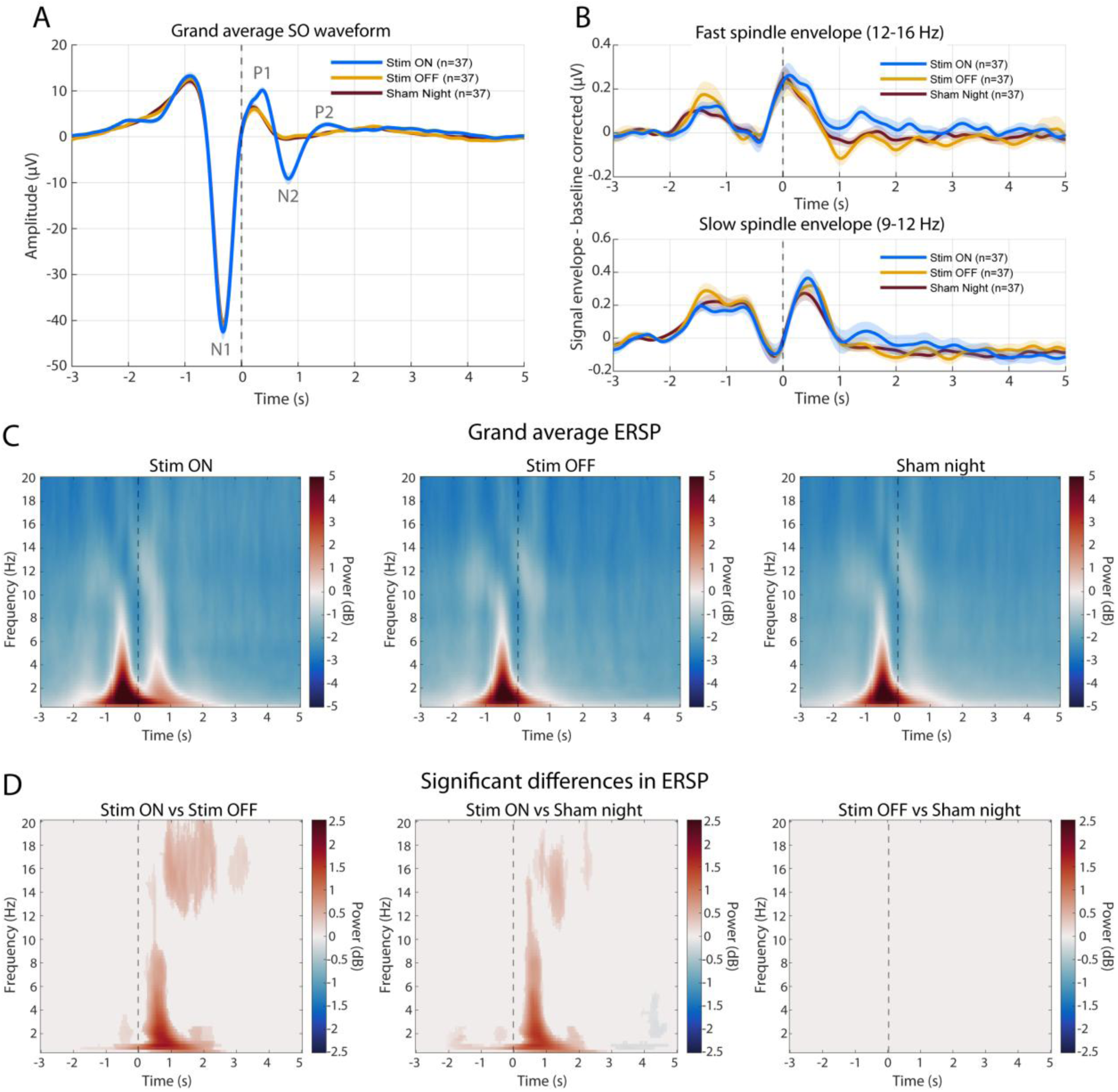
Acute electrophysiological effects of auditory stimulation. (A) Grand average waveform time-locked to online SO detection for Stim ON, Stim OFF, and Sham Night conditions. Labels indicate the detected SO negative peak (N1), the stimulation-induced positive peak (P1), secondary negative wave (N2), and subsequent positive rebound (P2). Shaded areas represent ±1 SEM. (B) Baseline-corrected signal envelope of fast (12–16 Hz, top) and slow (9–12 Hz, bottom) spindles, time-locked to SO detection. Shaded areas represent ±1 SEM. (C) Grand average event-related spectral perturbation (ERSP) for each condition, expressed in dB relative to a [−2.5, −2] s pre-detection baseline. (D) Significant pairwise differences in ERSP between conditions, assessed using cluster-based permutation testing (Monte Carlo, 1000 permutations, cluster-corrected p < 0.001). Only time-frequency pixels belonging to significant clusters are colored. Dashed vertical lines indicate SO detection time (t = 0). N = 37 for all conditions.

Event-related spectral perturbation analysis revealed the expected SO-locked power increase below 5 Hz across all conditions (Figure 3C). Notably, Stim ON additionally exhibited sustained post-stimulus power enhancement extending into the spindle frequency range. Quantitative spectral analyses confirmed significantly greater power in the SO, delta, theta, beta and spindle bands during Stim ON relative to both control conditions (Supplementary Figure 5). Pairwise significance maps (Figure 3D) demonstrated that these effects were restricted to the Stim ON vs Stim OFF and Stim ON vs Sham Night contrasts and localized to the [0, 2] s post-detection interval. No meaningful differences were observed between Stim OFF and Sham Night, supporting the electrophysiological equivalence of both sham conditions.

### 3.4 Associations between sleep electrophysiology and memory performance

Exploratory correlation analyses were performed to assess associations between stimulation-related electrophysiological responses and memory consolidation across tasks and retention intervals (Figure 4). Overall, no robust associations were observed between EEG metrics and overnight improvement in the word-pair task, despite this being the only behavioral outcome showing a significant stimulation effect at the group level.

**Figure 4.**
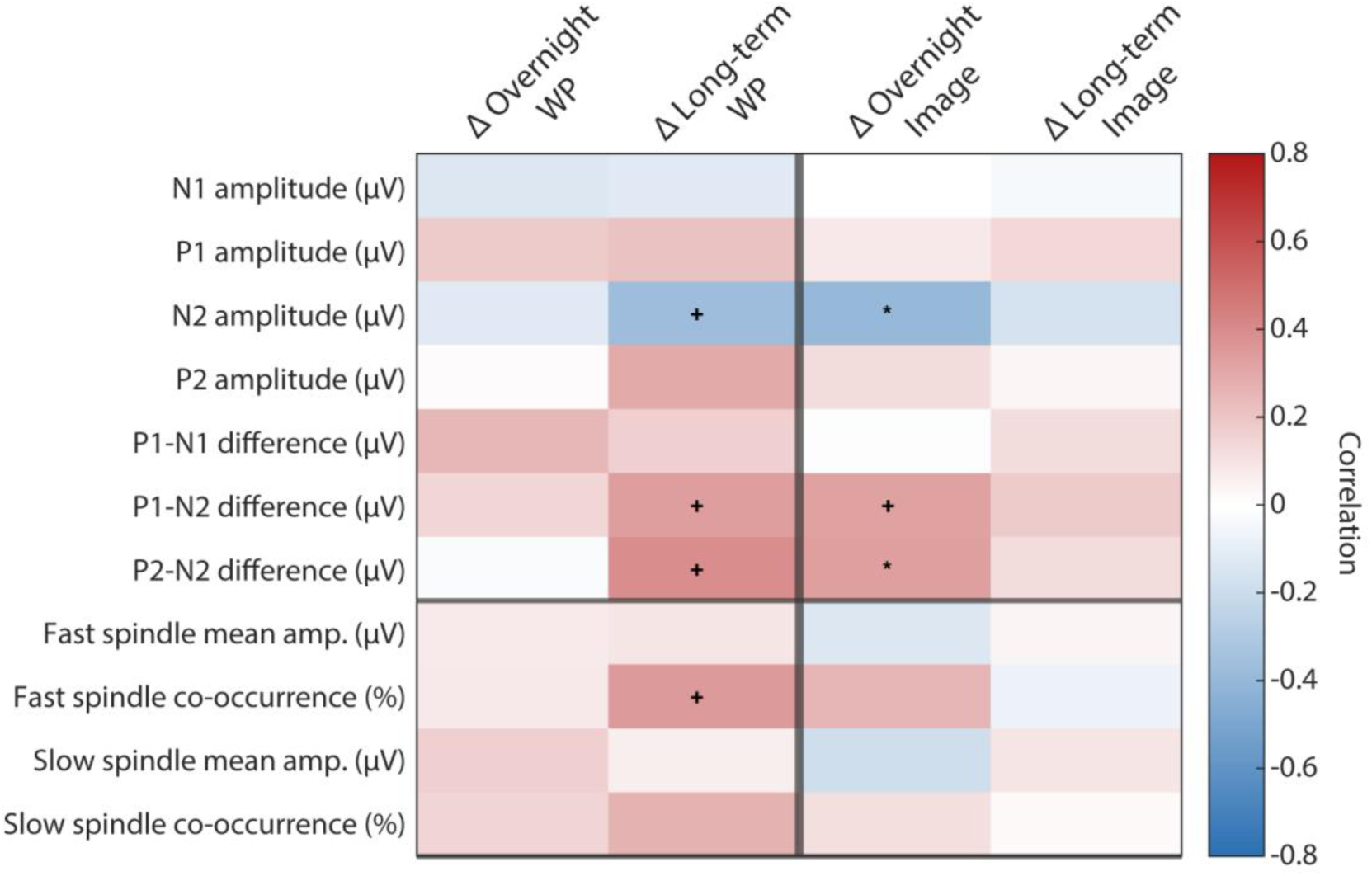
Correlation between stimulation-induced electrophysiological responses and memory consolidation. Heatmap of correlation coefficients linking EEG metrics and memory performance scores across tasks and retention intervals. EEG metrics are grouped into slow-wave amplitude descriptors (top) and spindle metrics (bottom). Memory outcomes are expressed as retention scores for the word-pair association task (WP) and visuospatial image recognition task (Image), at overnight and 7-day follow-up intervals. All analyses are exploratory and uncorrected for multiple comparisons. +p < 0.1; *p < 0.05.

However, a convergent pattern of associations emerged for several slow-wave amplitude metrics related to the stimulation-induced secondary slow wave, including the generated negative peak amplitude (N2), the P1–N2 peak-to-peak amplitude, and the N2–P2 peak-to-peak amplitude. These measures showed consistent associations with both long-term word-pair retention and overnight image recognition performance. Two of these associations reached nominal significance (*p* < 0.05), specifically for N2 amplitude and N2–P2 amplitude in relation to overnight image task performance, whereas the remaining related associations were trend-level (*p* < 0.1). In addition, fast spindle co-occurrence showed a trend-level association with long-term word-pair retention (*p* < 0.1).

### 3.5 Sleep structure

Sleep architecture did not differ significantly between stimulation and sham nights (Table 2). No significant effects of auditory stimulation were observed for total sleep time, sleep period time, sleep efficiency, wake after sleep onset, sleep onset latency, number of awakenings, or sleep stage distribution (all *p* > 0.05). These findings indicate that auditory stimulation did not measurably alter global sleep macrostructure.

**Table 2.** Sleep architecture metrics across experimental conditions. Values are mean ± SD; n = 37. The calibration night is shown for reference only; p-values correspond to paired comparisons between stimulation and sham nights (paired t-test or Wilcoxon signed-rank test depending on normality). SO coupling refers to the proportion of detected slow waves followed by a subsequent slow wave response. Abbreviations: N1/N2/N3, non-rapid eye movement sleep stages 1–3; REM, rapid eye movement; SOs, slow oscillations; SPT, sleep period time.

| | Calibration<br>night | Stim night | Sham night | Stim vs Sham<br>$p$-value |
| --- | --- | --- | --- | --- |
| Total sleep time [min] | 333.9 ± 79.7 | 353.5 ± 75.7 | 359.4 ± 74.8 | 0.59 |
| Sleep period time [min] | 426.9 ± 58.7 | 441.4 ± 59.7 | 434.1 ± 65.5 | 0.48 |
| Sleep efficiency [%] | 70.9 ± 16.3 | 73.7 ± 15.7 | 74.7 ± 15.1 | 0.65 |
| Wake after sleep onset [min] | 93.0 ± 70.9 | 87.9 ± 73.9 | 74.7 ± 58.5 | 0.21 |
| Sleep onset latency [min] | 39.3 ± 34.0 | 32.4 ± 36.6 | 35.6 ± 40.7 | 0.51 |
| Awakenings [#] | 22.3 ± 12.0 | 22.2 ± 9.2 | 21.9 ± 12.4 | 0.87 |
| Sleep stage changes [#] | 79.6 ± 34.5 | 82.6 ± 26.4 | 83.2 ± 36.5 | 0.91 |
| Stage Wake [min] | 138.2 ± 81.0 | 127.8 ± 79.0 | 122.6 ± 74.8 | 0.63 |
| Stage N1 [min] | 13.9 ± 9.0 | 14.1 ± 6.6 | 14.1 ± 8.4 | 0.99 |
| Stage N2 [min] | 256.6 ± 69.3 | 270.4 ± 58.1 | 271.0 ± 63.2 | 0.95 |
| Stage N3 [min] | 1.5 ± 6.3 | 1.8 ± 6.8 | 1.3 ± 6.2 | 0.64 |
| Stage REM [min] | 61.9 ± 28.7 | 67.1 ± 37.8 | 72.9 ± 36.7 | 0.30 |
| Stage Wake [% SPT] | 21.7 ± 16.1 | 19.3 ± 15.5 | 17.1 ± 12.6 | 0.31 |
| Stage N1 [% SPT] | 3.2 ± 2.0 | 3.2 ± 1.5 | 3.3 ± 1.9 | 0.75 |
| Stage N2 [% SPT] | 60.3 ± 14.5 | 61.7 ± 11.7 | 62.6 ± 11.8 | 0.63 |
| Stage N3 [% SPT] | 0.4 ± 1.5 | 0.4 ± 1.4 | 0.3 ± 1.4 | 0.70 |
| Stage REM [% SPT] | 14.5 ± 6.3 | 15.4 ± 8.5 | 16.7 ± 8.0 | 0.29 |
| SOs per minute [#] | 1.9 ± 1.5 | 3.5 ± 1.5 | 3.9 ± 1.9 | 0.18 |
| SOs - Detected [#] | 448.4 ± 488.7 | 859.5 ± 584.4 | 900.9 ± 462.9 | 0.60 |
| SOs - Stimulated [#] | 228.8 ± 250.6 | 437.5 ± 295.8 | 0.0 ± 0.0 | |
| SOs - Sham [#] | 219.6 ± 238.3 | 422.0 ± 289.0 | 900.9 ± 462.9 | |
| SOs - Coupled [%] | 6.2 ± 5.1 | 10.2 ± 5.5 | 6.7 ± 2.9 | <0.001 |
| Coupled SOs after Stim [%] | 8.7 ± 6.3 | 14.2 ± 7.9 | 0.0 ± 0.0 | |
| Coupled SOs after Sham [%] | 3.7 ± 3.8 | 6.0 ± 3.0 | 6.7 ± 2.9 | |

The total number of detected SOs, as well as the rate of SOs per minute, was numerically higher during sham nights compared with stimulation nights, although these differences did not reach statistical significance (Table 2). In contrast, the proportion of detected SOs followed by a subsequent slow oscillation response (coupled SOs) was significantly greater during stimulation nights (*p* < 0.001). This effect was primarily driven by SOs receiving auditory stimulation, whereas coupling rates for non-stimulated SOs during stimulation nights were comparable to those observed during sham nights.

### 3.6 Subjective measures and experimental control analyses

No significant differences were observed between stimulation and sham nights for any subjective sleep quality or sleepiness measure (all *p* > 0.05; Table 3). SF-A/R scores indicated comparable subjective sleep experience across conditions, including perceived sleep quality, difficulties initiating or maintaining sleep, feeling of recovery after sleep, and psychosomatic symptoms during the night. Likewise, Stanford Sleepiness Scale scores before sleep onset and after waking did not differ between conditions.

**Table 3.**
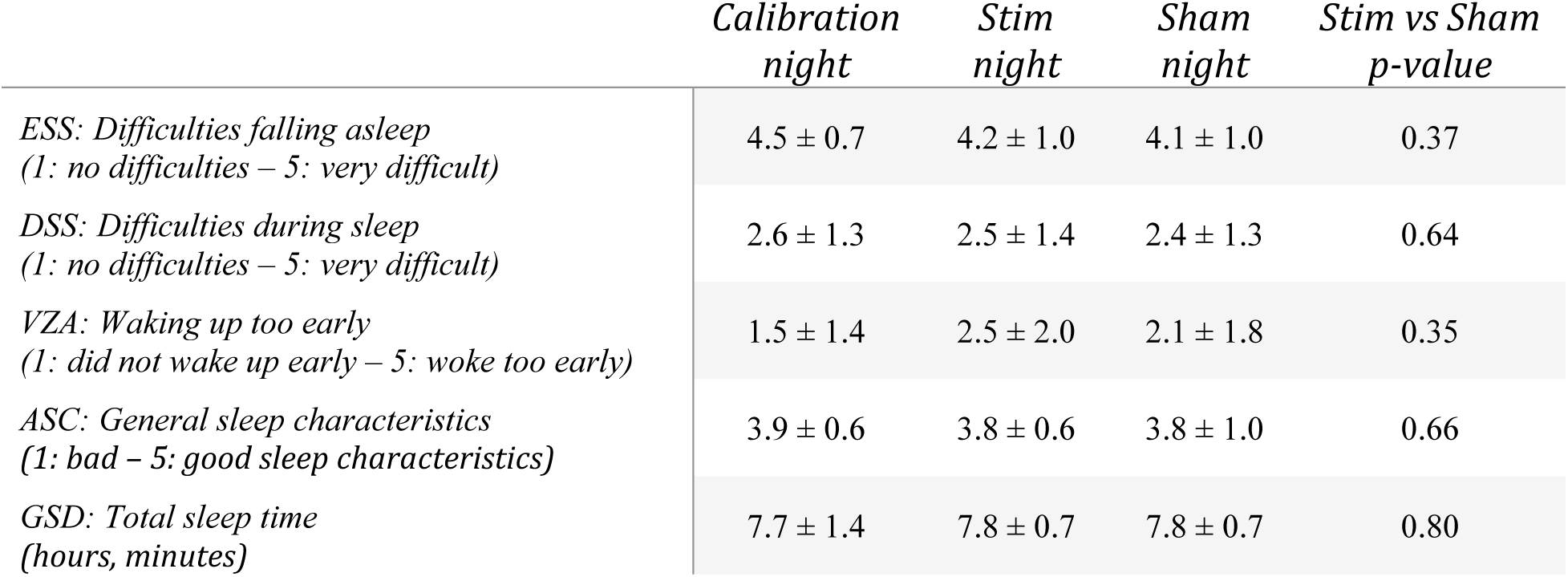
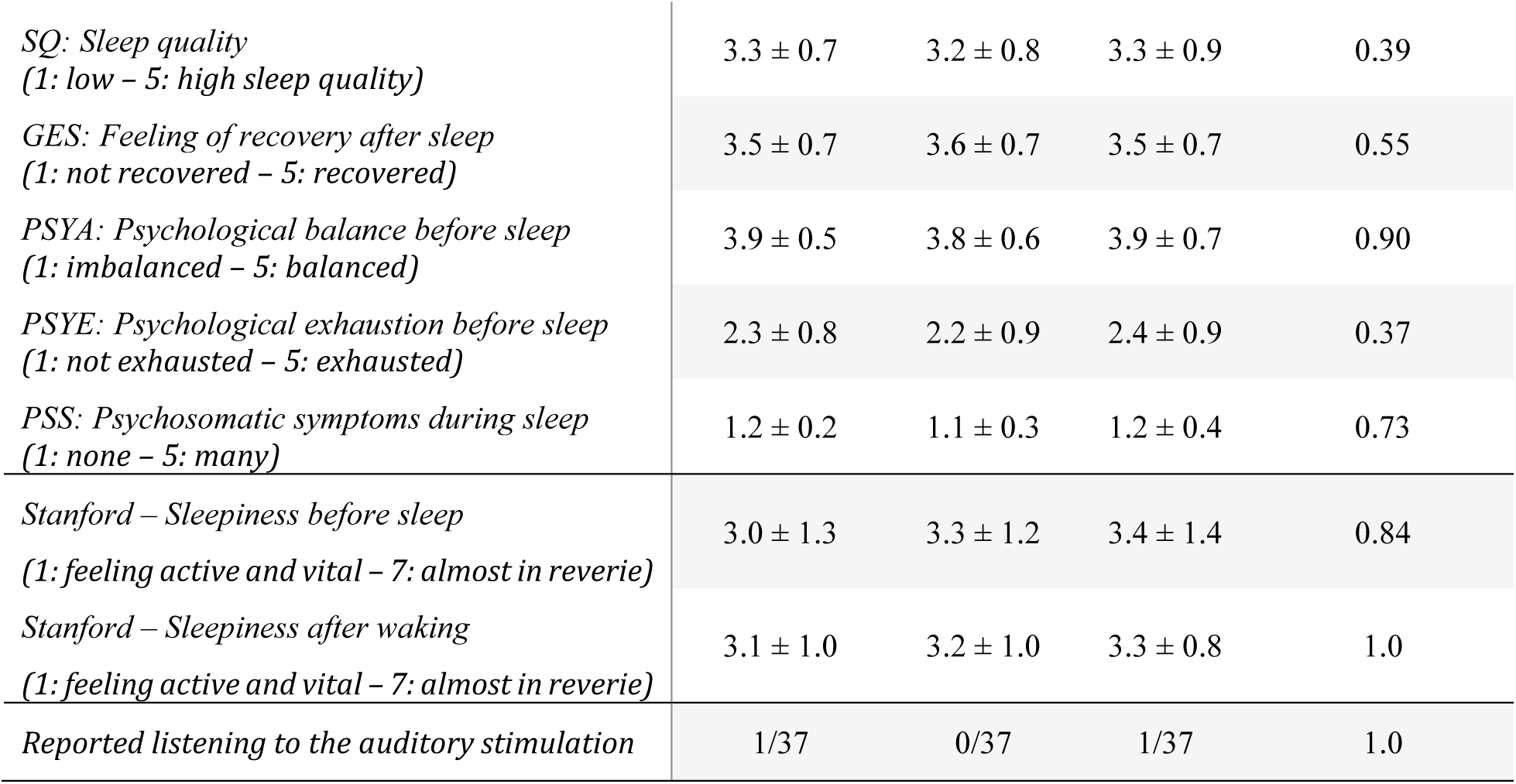
Subjective sleep metrics across experimental conditions. Values are mean ± SD; n = 37. The calibration night is shown for reference only; p-values correspond to paired comparisons between stimulation and sham nights (paired t-test or Wilcoxon signed-rank test depending on normality).

| | Calibration<br>night | Stim<br>night | Sham<br>night | Stim vs Sham<br>$p$-value |
| --- | --- | --- | --- | --- |
| ESS: Difficulties falling asleep<br>(1: no difficulties – 5: very difficult) | 4.5 ± 0.7 | 4.2 ± 1.0 | 4.1 ± 1.0 | 0.37 |
| DSS: Difficulties during sleep<br>(1: no difficulties – 5: very difficult) | 2.6 ± 1.3 | 2.5 ± 1.4 | 2.4 ± 1.3 | 0.64 |
| VZA: Waking up too early<br>(1: did not wake up early – 5: woke too early) | 1.5 ± 1.4 | 2.5 ± 2.0 | 2.1 ± 1.8 | 0.35 |
| ASC: General sleep characteristics<br>(1: bad – 5: good sleep characteristics) | 3.9 ± 0.6 | 3.8 ± 0.6 | 3.8 ± 1.0 | 0.66 |
| GSD: Total sleep time<br>(hours, minutes) | 7.7 ± 1.4 | 7.8 ± 0.7 | 7.8 ± 0.7 | 0.80 |
| *SQ: Sleep quality*<br>(1: low – 5: high sleep quality) | 3.3 ± 0.7 | 3.2 ± 0.8 | 3.3 ± 0.9 | 0.39 |
| *GES: Feeling of recovery after sleep*<br>(1: not recovered – 5: recovered) | 3.5 ± 0.7 | 3.6 ± 0.7 | 3.5 ± 0.7 | 0.55 |
| *PSYA: Psychological balance before sleep*<br>(1: imbalanced – 5: balanced) | 3.9 ± 0.5 | 3.8 ± 0.6 | 3.9 ± 0.7 | 0.90 |
| *PSYE: Psychological exhaustion before sleep*<br>(1: not exhausted – 5: exhausted) | 2.3 ± 0.8 | 2.2 ± 0.9 | 2.4 ± 0.9 | 0.37 |
| *PSS: Psychosomatic symptoms during sleep*<br>(1: none – 5: many) | 1.2 ± 0.2 | 1.1 ± 0.3 | 1.2 ± 0.4 | 0.73 |
| *Stanford – Sleepiness before sleep*<br>(1: feeling active and vital – 7: almost in reverie) | 3.0 ± 1.3 | 3.3 ± 1.2 | 3.4 ± 1.4 | 0.84 |
| *Stanford – Sleepiness after waking*<br>(1: feeling active and vital – 7: almost in reverie) | 3.1 ± 1.0 | 3.2 ± 1.0 | 3.3 ± 0.8 | 1.0 |
| *Reported listening to the auditory stimulation* | 1/37 | 0/37 | 1/37 | 1.0 |

Only two participants reported perceiving auditory stimulation during any study session, one during the calibration night and one during the sham night. No participants reported hearing stimulation during the experimental night that included stimulation, supporting successful auditory masking. Environmental conditions remained stable across sessions, with comparable temperature and humidity values during calibration, stimulation, and sham nights (Supplementary Table 1).

## 4 Discussion

This study provides the first double-blind, placebo-controlled evidence that a single night of closed-loop auditory stimulation (CLAS) can significantly improve overnight declarative memory consolidation in patients with biomarker-confirmed AD at an early stage with an amnestic clinical phenotype. The observed effect on declarative memory (Cohen’s d_z_ = 0.42) falls within the range reported in previous auditory stimulation studies and is particularly notable given the smaller and more variable effect sizes typically reported in older populations [16,37].

### 4.1 Mechanistic validation of stimulation effects

Auditory stimulation produced robust electrophysiological effects. At the waveform level, stimulation elicited the characteristic sequence of an enhanced positive SO up-state, accompanied by increased fast spindle activity, followed by a secondary SO cycle [15,38]. These effects were reflected both in the grand-average waveforms and in the quantitative analyses, which showed significant increases in the stimulation-induced slow wave peak amplitudes. In parallel, stimulation significantly enhanced fast spindle activity, both in terms of spindle amplitude and SO-spindle co-occurrence, whereas no significant effects were observed for slow spindles.

The observed effects closely resemble the canonical electrophysiological response previously reported in healthy younger and older adults undergoing phase-locked auditory stimulation [15,18,20,21,38]. The enhancement of fast spindle activity is particularly relevant given the proposed role of SO-spindle coupling in coordinating hippocampal-neocortical communication during memory consolidation [6,7,39] and their relationship with degeneration in AD [40]. Together, these findings indicate that the cortical and thalamocortical circuits engaged by CLAS remain recruitable in patients with early symptomatic AD despite ongoing neurodegenerative pathology [11,13].

Moreover, the absence of significant differences between Stim OFF and the Sham night across waveform, spindle, and spectral analyses provides strong support for the internal validity of the protocol [41]. Because unstimulated slow oscillations on the stimulation night did not differ from spontaneous slow oscillations recorded during the sham condition, the observed enhancements can be attributed specifically to the auditory stimulation rather than to nonspecific differences in sleep state, night-to-night variability, or experimental context.

### 4.2 Effects on memory consolidation: specificity and boundaries

CLAS selectively improved overnight retention of word-pair associations, with no significant effects on visuospatial image recognition or working memory. This pattern is consistent with the literature, where word-pair associative learning has been the most frequently targeted and most consistently improved memory domain [15,17,42–45]. By contrast, evidence for behavioral benefits in visuospatial paradigms remains limited, and null findings have been reported despite reliable enhancement of slow oscillations and spindles [44]. A plausible explanation is that these tasks differ in their dependence on hippocampal processes supported by slow oscillations and sleep spindles [46]. Associative word-pair learning depends critically on hippocampal binding and is therefore particularly sensitive to sleep-dependent consolidation [6,7], whereas recognition-based visuospatial tasks likely rely more strongly on familiarity-related processes supported by distributed cortical networks [47–49]. Working memory, in turn, depends primarily on short-term maintenance and executive control rather than on the consolidation mechanisms targeted by CLAS. The selective benefit observed here is therefore consistent with models assigning a central role to slow oscillation-mediated hippocampal-neocortical communication in associative memory consolidation [6,7,50,51]. At the same time, the word-pair literature is not uniform, as several studies have failed to replicate the original benefit, particularly in older samples or with semantically incongruent materials [38,52,53].

Interpretation of the null result for image recognition is further limited by a methodological limitation of the study. The order of the stimulus sets was fixed across participants, and image recognition—but not word-pair learning—showed a significant night-order effect, suggesting that the two image sets may have differed in difficulty. Such an imbalance would increase performance variability and reduce sensitivity to detect a genuine stimulation effect [54]. Consequently, the absence of an image-recognition benefit cannot be attributed unambiguously either to differences in task-related sleep dependence or to stimulus-set characteristics. Future studies should therefore verify psychometric equivalence of visuospatial stimulus sets before deployment in crossover designs.

The overnight benefit observed for word-pair learning was not sustained at the seven-day follow-up. A single night of stimulation may be sufficient to influence short-term consolidation without necessarily producing memory traces robust enough to withstand subsequent days of interference [13]. In addition, the long-term analysis was affected by technical data loss, which reduced statistical power to detect potentially smaller delayed effects. These findings indicate that sleep-dependent consolidation mechanisms remain modifiable in AD patients with early-stage amnestic impairment, while leaving open whether repeated stimulation is required to produce more durable cognitive benefits, as suggested by emerging multi-night intervention studies [18,21].

### 4.3 Brain-behavior associations and candidate response biomarkers

Exploratory correlation analyses revealed that the amplitude of the stimulation-evoked secondary slow-wave was associated with both overnight image recognition and long-term word-pair retention. Notably, this association was absent for the overnight word-pair outcome, the only measure showing a significant group-level benefit of stimulation.

The presence of convergent associations across two distinct memory outcomes suggests that the observed electrophysiological responses are not arbitrary. Measures describing the stimulation-evoked secondary slow-wave response emerged as the strongest correlates of memory performance, consistent with previous reports linking stimulation-induced enhancement of slow oscillations or spindle activity to behavioral improvement following auditory sleep stimulation [15,18,20,21]. Together, these observations suggest that CLAS engaged neural mechanisms relevant to memory processing. Nevertheless, given the exploratory nature of the analyses and the absence of correction for multiple comparisons, these findings should be interpreted cautiously.

The absence of a significant correlation for the overnight word-pair benefit, however, differs from prior studies. One relevant factor contributing to this discrepancy may be the heterogeneity across patients in disease burden, sleep physiology, and cognitive reserve, all of which may shape the relationship between neural responsiveness and behavioral outcome.

Although participants were recruited according to an amnestic MCI clinical phenotype, baseline cognitive performance varied considerably across the cohort. Thus, the findings should be interpreted as applying to an early AD population characterized by an amnestic MCI clinical phenotype but with heterogeneous cognitive severity. Still, the absence of a detectable correlation should not be interpreted as evidence against a mechanistic contribution of sleep-dependent consolidation processes, particularly given the modest sample size available for these exploratory analyses.

A growing body of work indicates substantial inter-individual variability in responsiveness to auditory sleep stimulation, particularly in older adults and populations with cognitive impairment [19,20,22,23]. In this context, the stimulation-evoked oscillatory profile may represent a candidate biomarker of treatment responsiveness, analogous to the use of EEG-derived measures for AD stratification [55]. Future studies should determine whether such electrophysiological markers can identify patients most likely to benefit from CLAS and thereby support individualized stimulation protocols [11,56].

### 4.4 Safety, tolerability, and preservation of sleep architecture

A prerequisite for the clinical applicability of auditory sleep stimulation is that it enhances sleep-related neural processes without disrupting sleep itself. In the present study, stimulation produced no detectable alterations in sleep macrostructure or perceived sleep quality, supporting its suitability for patients with AD, a population particularly vulnerable to sleep fragmentation and nocturnal disturbances [9,57].

An additional strength of the protocol was the successful maintenance of blinding. Only one patient across all experimental nights reported perceiving auditory stimulation, and that report occurred during the sham night. This absence of conscious perception indicates that stimulation was delivered below the threshold of awareness while remaining sufficient to induce robust electrophysiological responses. These results support the clinical acceptability of the intervention and strengthen the interpretation that the observed memory benefits resulted from modulation of sleep-dependent neural processes rather than nonspecific behavioral or expectancy effects.

### 4.5 Limitations and future directions

Several limitations should be considered when interpreting these findings. First, the intervention involved only a single night of stimulation. Although this design allows establishing causal effects on sleep physiology and overnight memory consolidation [15], it does not address whether repeated stimulation can produce cumulative cognitive benefits or influence disease-related biological processes. Emerging evidence from multi-night protocols suggests that longer interventions may be required to achieve durable effects on memory and potentially on AD-related biomarkers [18,19,21].

Second, the study was not powered to detect small effects on long-term memory retention. Technical difficulties affecting the long-term word-pair assessment reduced the available sample size for this analysis and may have limited sensitivity to detect delayed cognitive benefits. The absence of significant effects at the one-week follow-up should therefore be interpreted cautiously.

Finally, although participants were selected based on both clinical and biomarker evidence of AD pathology, molecular outcome measures were not included. Given the relationship between sleep, amyloid clearance, tau pathology, and neurodegeneration [9,11,58], future trials should incorporate biomarker endpoints alongside cognitive outcomes to assess how CLAS influences disease-related processes [18,21].

Overall, the present study provides rigorous placebo-controlled evidence that sleep-dependent memory consolidation remains amenable to modulation through closed-loop auditory stimulation in patients with biomarker-confirmed early symptomatic AD and an amnestic clinical phenotype. The robust electrophysiological responses and favorable tolerability profile support further evaluation of this approach in larger studies. Future work should prioritize multi-night home-based interventions, integration of biomarker outcomes, and prospective validation of physiological markers of treatment responsiveness to determine the long-term clinical potential of sleep-based neuromodulation across the AD continuum.

## Data Availability

The data supporting the findings of the study can be available from the authors upon reasonable request.

## Acknowledgments

This study has been funded with grants by the Horizon-EIC-2022 call (BAYFLEX: 101099555), the Horizon-RIA-2023 call (MANOLO: 101135782), the Proyecto Estratégico para la Recuperación y Transformación Económica (PERTE) para la Salud de Vanguardia (Multi-País: EXP-00170833/PAIS-20241086), the Diputación General de Aragón -Subvenciones I+D Movilidad sostenible y sector farmacéutico (ORDEN EPE/676/2023, expediente IDMF/2023/0007), and the Spanish Ministry of Science, Innovation and Universities through the Torres Quevedo 2022 contract scheme (PTQ2022-012518, E.J-G.) and the Doctorados Industriales 2024 program (DIN2024-014259-2, F.R.). A.P-L. is partly supported by the Healthy Aging Initiative, the Eleanor and Herbert Bearak Memory Wellness for Life Program, Diane and Mark Goldman, and grants from the National Institutes of Health (R01AG076708; R01AG059089), Jack Satter Foundation, and BrightFocus Foundation.

## CRediT authorship contribution statement

**E.L-L.**: Conceptualization, Data Curation, Formal analysis, Investigation, Methodology, Project administration, Software, Supervision, Validation, Visualization, Writing - Original Draft. **D.O.**: Data Curation, Formal analysis, Investigation, Project administration, Validation, Visualization, Writing - Original Draft. **G.F.**: Data Curation, Investigation, Project administration, Validation, Visualization, Writing - Review & Editing. **E.J-G.**: Conceptualization, Methodology, Project administration, Supervision, Writing - Original Draft. **J.G.K.**: Conceptualization, Methodology, Software, Supervision, Validation, Writing - Review & Editing. **M.S-T.**: Data Curation, Investigation, Methodology, Software, Validation, Writing - Review & Editing. **E.H-P.**: Data Curation, Investigation, Methodology, Software, Validation, Writing - Review & Editing. **C.E.**: Methodology, Software, Validation, Writing - Review & Editing. **J.S-J.**: Investigation, Methodology, Project administration, Visualization, Writing - Review & Editing. **F.R.**: Investigation, Methodology, Project administration, Writing - Review & Editing. **L.M.**: Funding acquisition, Methodology, Supervision, Writing - Review & Editing. **C.M.L.**: Conceptualization, Resources, Writing - Review & Editing. **G.P.**: Supervision, Writing - Review & Editing. **A.P-L.**: Supervision, Writing - Review & Editing. **E.M.M.**: Conceptualization, Resources, Supervision, Writing - Review & Editing. **E.M-F.**: Conceptualization, Resources, Supervision, Writing - Review & Editing. **J.M.**: Conceptualization, Funding acquisition, Supervision, Writing - Review & Editing.

## Supplementary materials

**Supplementary Figure 1.**
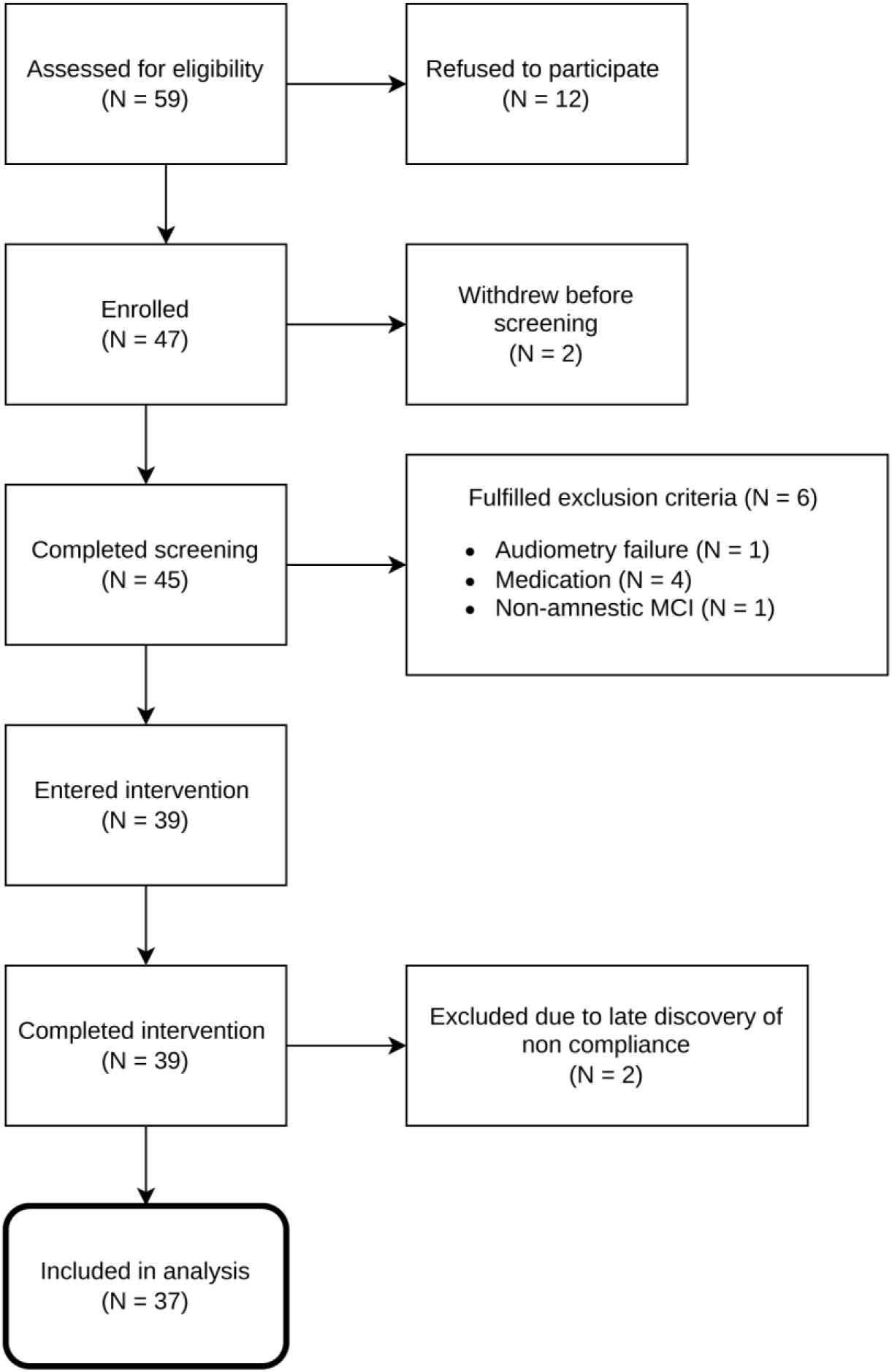
Flow diagram of patient recruitment. Of 59 patients assessed for eligibility, a total of 39 entered and completed the intervention phase. Two were subsequently excluded due to late discovery of protocol non-compliance, resulting in a final analyzed sample of N = 37.

**Supplementary Figure 2.**
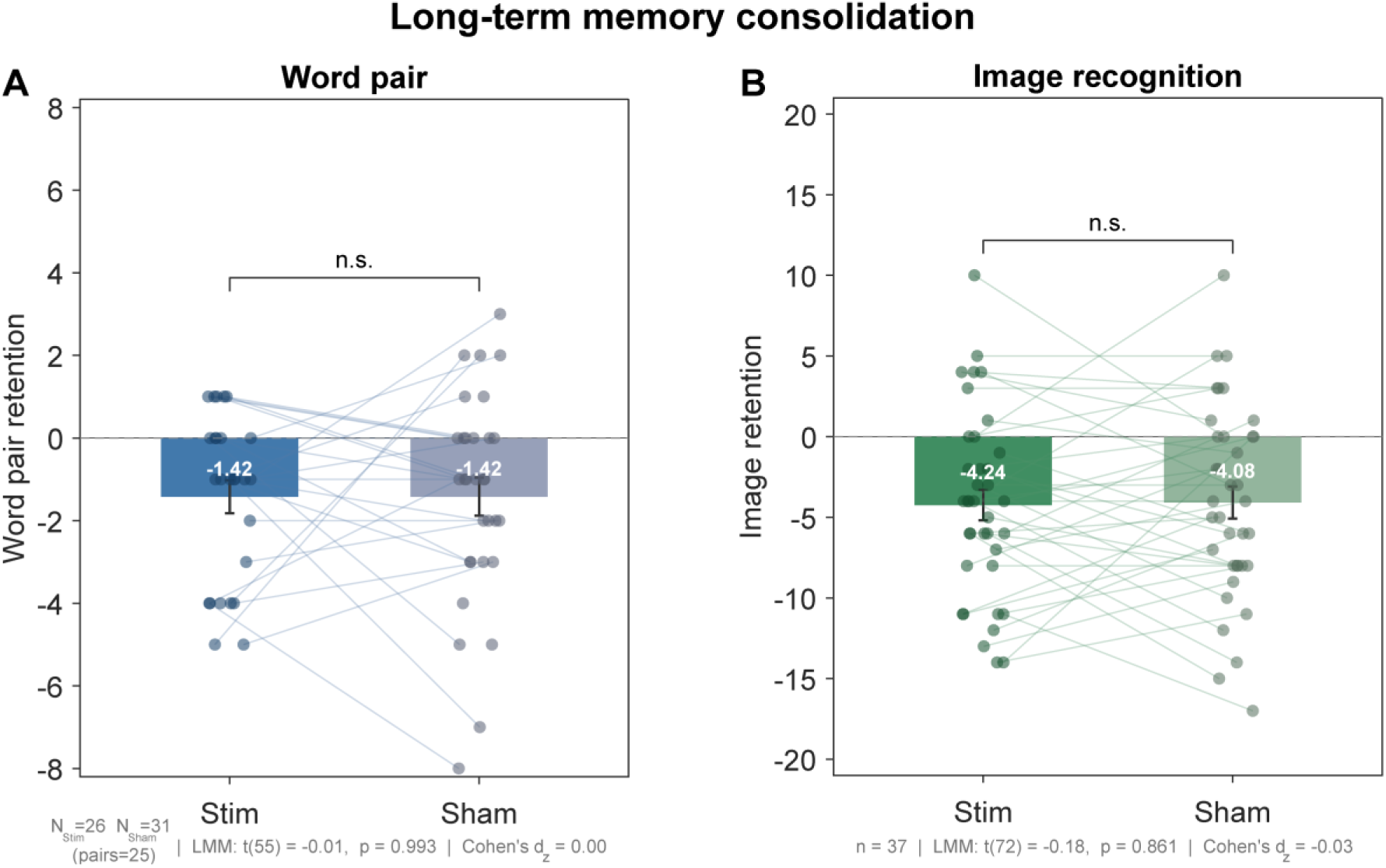
Effects of CLAS on long-term memory consolidation. No significant difference in long-term consolidation (LT − immediate recall) was detected between the stimulation and sham conditions for either the word-pair (A) or visuospatial image recognition (B) task. n.s., not significant.

**Supplementary Figure 3.**
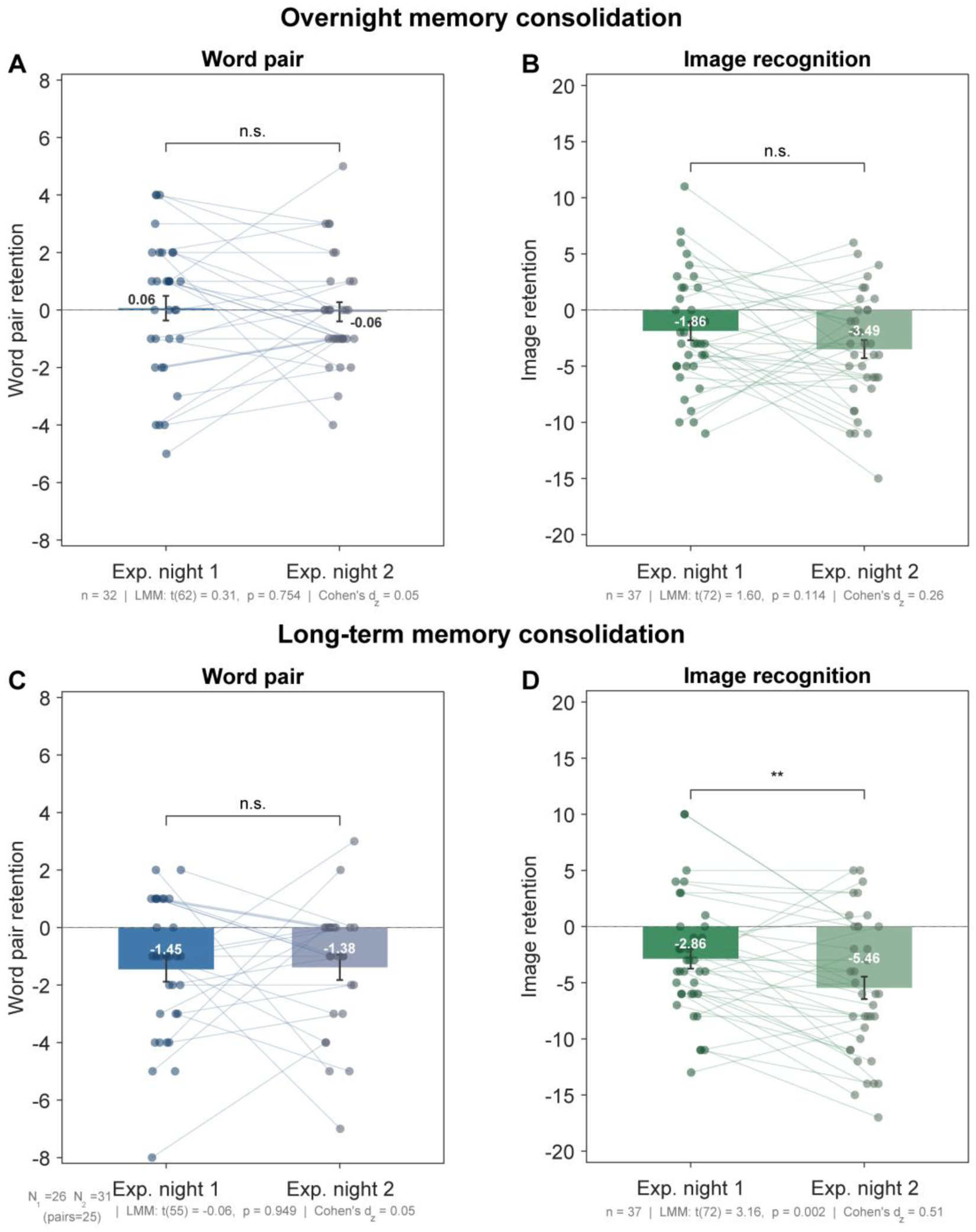
Influence of experimental night order on memory retention scores. Memory retention was compared between the first and second experimental nights, irrespective of condition assignment. Overnight consolidation (morning − immediate recall) showed no significant night-order effect for the word-pair (A) or visuospatial image recognition (B) task. Long-term consolidation (LT − immediate recall) showed no effect for the word-pair task (C), whereas image recognition (D) exhibited significantly greater forgetting on the second night. n.s., not significant; **p < 0.01

**Supplementary Figure 4.**
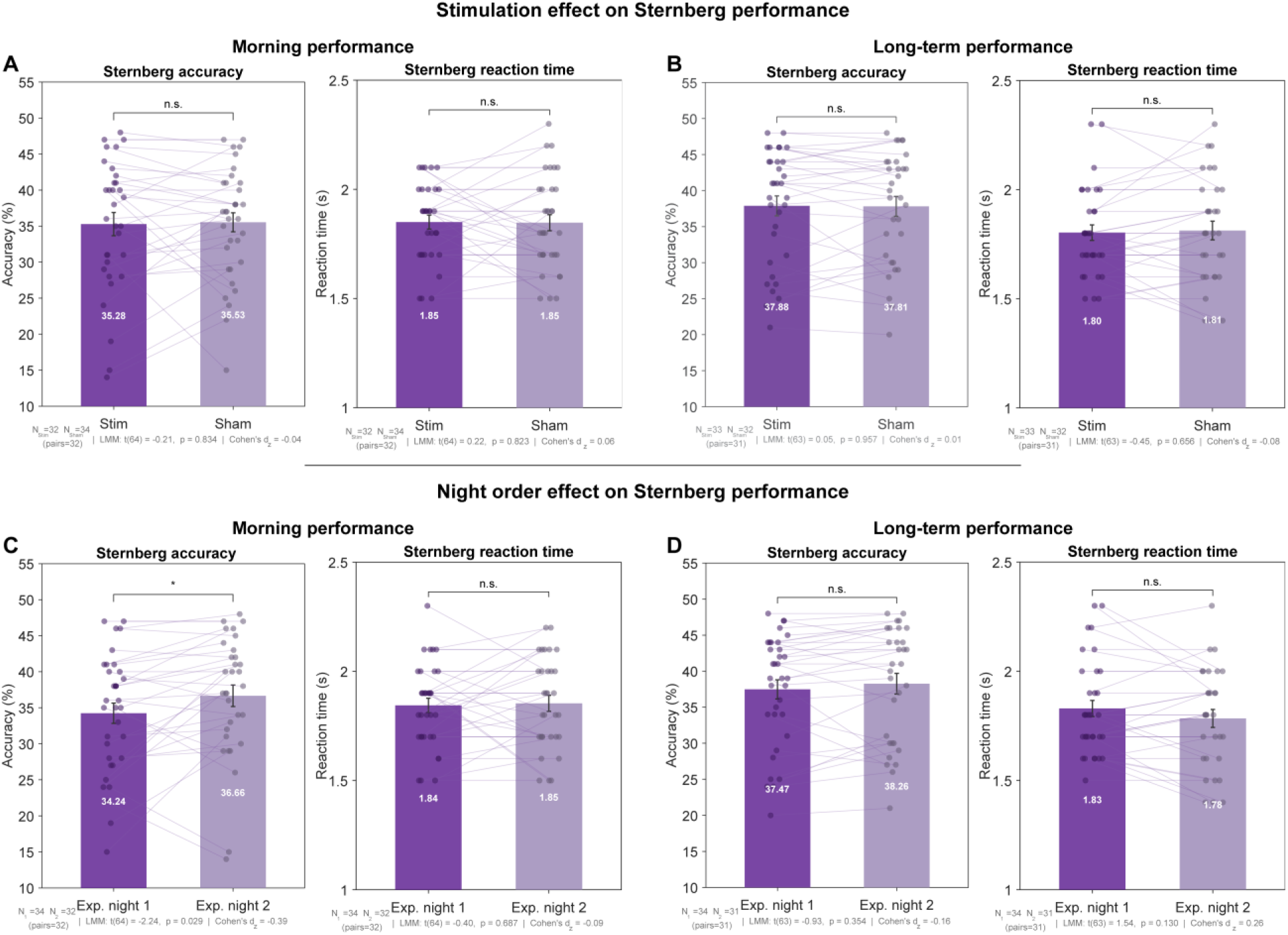
Effects of auditory stimulation and experimental night order on working memory performance. No significant differences in Sternberg task performance (accuracy and reaction time) were detected between stimulation and sham conditions at either morning (A) or long-term follow-up (B) assessments. Comparison between the first and second experimental nights, irrespective of condition assignment, revealed higher accuracy on the second night at the morning assessment (C), whereas no other night-order effects were observed (D). Displayed p-values are uncorrected; the night-order effect on morning accuracy did not survive correction for multiple comparisons. n.s., not significant; *p < 0.05.

**Supplementary Figure 5.**
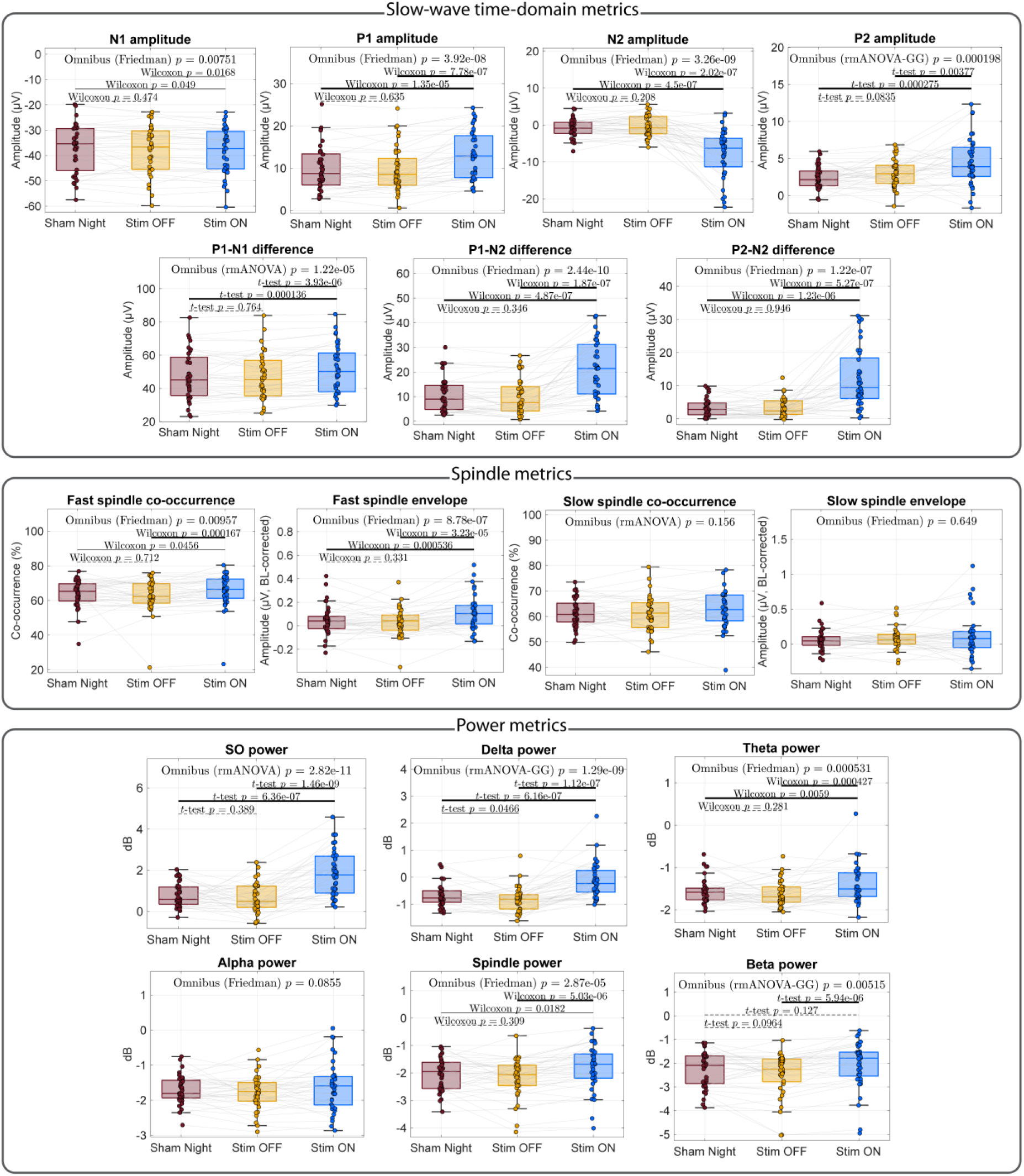
Quantitative electrophysiological metrics across stimulation conditions. Boxplots showing group-level distributions for Sham Night, Stim OFF, and Stim ON conditions across three metric categories. Top: Slow-wave time-domain metrics, including peak amplitudes of the detected slow wave (N1), the stimulation-induced positive peak (P1), the secondary negative wave (N2), and the subsequent positive rebound (P2), together with peak-to-peak difference metrics (P1−N1, P1−N2, P2−N2). Middle: Spindle metrics, including co-occurrence rate and baseline-corrected envelope amplitude for fast (12–16 Hz) and slow (9–12 Hz) spindles. Bottom: Baseline-corrected spectral power in six frequency bands (SO, 0.5–1 Hz; delta, 1–4 Hz; theta, 4–8 Hz; alpha, 8–12 Hz; spindle, 12–16 Hz; beta, 15–30 Hz), averaged over the [0, 2] s post-detection window. For each metric, the omnibus test result (rmANOVA or Friedman, depending on normality) is shown, followed by pairwise post-hoc comparisons (paired t-test or Wilcoxon signed-rank test). Displayed p-values are uncorrected. Horizontal lines connecting pairwise comparisons are thick solid for Bonferroni-corrected p-values < 0.05, thin solid for uncorrected p-values < 0.05 that did not survive correction, and thin dashed for non-significant comparisons (p ≥ 0.05). Individual data points represent single patients. Abbreviations: BL, baseline; GG, Greenhouse-Geisser correction.

**Supplementary Table 1.**
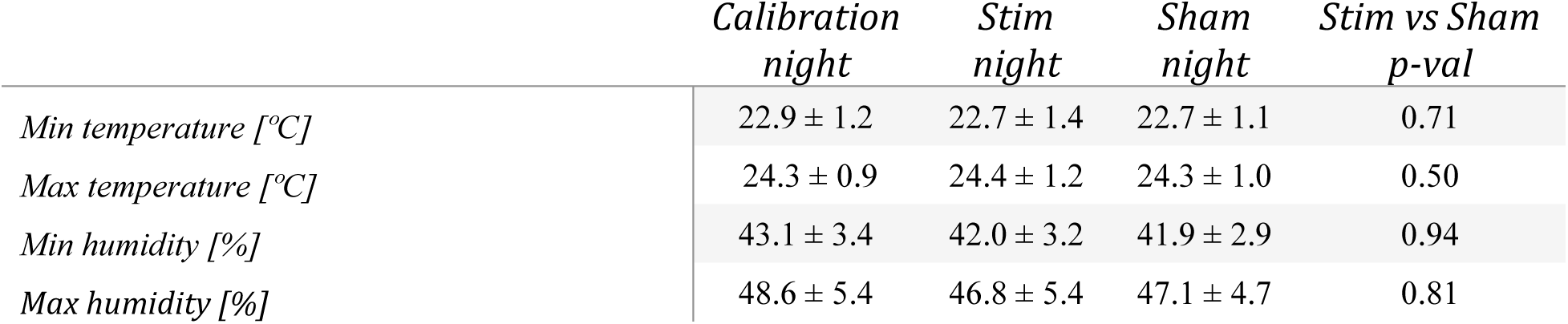
Environmental data of the sleep laboratory. Values are presented as mean ± SD. P-values correspond to comparisons between stimulation and sham nights

## References

[1] GBD 2019 Dementia Forecasting Collaborators (2022). Estimation of the global prevalence of dementia in 2019 and forecasted prevalence in 2050: an analysis for the Global Burden of Disease Study 2019. The Lancet Public health. 10.1016/S2468-2667(21)00249-8.

[2] Jahn, H. (2013). Memory loss in Alzheimer’s disease. Dialogues in Clinical Neuroscience. 10.31887/DCNS.2013.15.4/hjahn.

[3] Morris, R.G. and Kopelman, M.D. (1986). The Memory Deficits in Alzheimer-type Dementia: A Review. The Quarterly Journal of Experimental Psychology. 10.1080/14640748608401615.

[4] Sperling, R.A. et al. (2010). Functional Alterations in Memory Networks in Early Alzheimer’s Disease. NeuroMolecular Medicine. 10.1007/s12017-009-8109-7.

[5] Rasch, B. and Born, J. (2013). About Sleep’s Role in Memory. Physiological Reviews. 10.1152/physrev.00032.2012.

[6] Klinzing, J.G. et al. (2019). Mechanisms of systems memory consolidation during sleep. Nature Neuroscience. 10.1038/s41593-019-0467-3.

[7] Diekelmann, S. and Born, J. (2010). The memory function of sleep. Nature Reviews Neuroscience. 10.1038/nrn2762.

[8] Bubu, O.M. et al. (2017). Sleep, Cognitive impairment, and Alzheimer’s disease: A Systematic Review and Meta-Analysis. Sleep. 10.1093/sleep/zsw032.

[9] Ju, Y.-E.S. et al. (2014). Sleep and Alzheimer disease pathology-a bidirectional relationship. Nature Reviews Neurology. 10.1038/nrneurol.2013.269.

[10] Mander, B.A. et al. (2015). β-amyloid disrupts human NREM slow waves and related hippocampus-dependent memory consolidation. Nature Neuroscience. 10.1038/nn.4035.

[11] Mander, B.A. et al. (2016). Sleep: A Novel Mechanistic Pathway, Biomarker, and Treatment Target in the Pathology of Alzheimer’s Disease? Trends in neurosciences. 10.1016/j.tins.2016.05.002.

[12] Wunderlin, M. et al. (2020). The role of slow wave sleep in the development of dementia and its potential for preventative interventions. Psychiatry Research: Neuroimaging. 10.1016/j.pscychresns.2020.111178.

[13] Hanert, A. et al. (2024). Reduced overnight memory consolidation and associated alterations in sleep spindles and slow oscillations in early Alzheimer’s disease. Neurobiology of Disease. 10.1016/J.NBD.2023.106378.

[14] Sharon, O. et al. (2025). Slow wave synchrony during NREM sleep tracks cognitive impairment in prodromal Alzheimer’s disease. Alzheimer’s and Dementia. 10.1002/alz.70247.

[15] Ngo, H.-V.V. et al. (2013). Auditory Closed-Loop Stimulation of the Sleep Slow Oscillation Enhances Memory. Neuron. 10.1016/J.NEURON.2013.03.006.

[16] Wunderlin, M. et al. (2021). Modulating overnight memory consolidation by acoustic stimulation during slow-wave sleep: a systematic review and meta-analysis. Sleep. 10.1093/sleep/zsaa296.

[17] Papalambros, N.A. et al. (2017). Acoustic Enhancement of Sleep Slow Oscillations and Concomitant Memory Improvement in Older Adults. Frontiers in Human Neuroscience. 10.3389/fnhum.2017.00109.

[18] Wunderlin, M. et al. (2023). Acoustic stimulation during sleep predicts long-lasting increases in memory performance and beneficial amyloid response in older adults. Age and Ageing. 10.1093/ageing/afad228.

[19] Lustenberger, C. et al. (2022). Auditory deep sleep stimulation in older adults at home: a randomized crossover trial. Communications Medicine. 10.1038/s43856-022-00096-6.

[20] Papalambros, N.A. et al. (2019). Acoustic enhancement of sleep slow oscillations in mild cognitive impairment. Annals of Clinical and Translational Neurology. 10.1002/acn3.796.

[21] Zeller, C.J. et al. (2024). Multi-night acoustic stimulation is associated with better sleep, amyloid dynamics, and memory in older adults with cognitive impairment. GeroScience. 10.1007/s11357-024-01195-z.

[22] Van den Bulcke, L., et al. (2023). Acoustic stimulation as a promising technique to enhance slow-wave sleep in Alzheimer’s disease: results of a pilot study. Journal of Clinical Sleep Medicine. 10.5664/jcsm.10778.

[23] Van den Bulcke, L., et al. (2025). Acoustic Stimulation to Improve Slow-Wave Sleep in Alzheimer’s Disease: A Multiple Night At-Home Intervention. American Journal of Geriatric Psychiatry. 10.1016/j.jagp.2024.07.002.

[24] Pocock, S.J. and Simon, R. (1975). Sequential Treatment Assignment with Balancing for Prognostic Factors in the Controlled Clinical Trial. Biometrics. 10.2307/2529712.

[25] Jack, C.R. et al. (2024). Revised criteria for diagnosis and staging of Alzheimer’s disease: Alzheimer’s Association Workgroup. Alzheimer’s & Dementia. 10.1002/alz.13859.

[26] Dubois, B. et al. (2024). Alzheimer Disease as a Clinical-Biological Construct-An International Working Group Recommendation. JAMA neurology. 10.1001/jamaneurol.2024.3770.

[27] Faul, F. et al. (2007). G*Power 3: A flexible statistical power analysis program for the social, behavioral, and biomedical sciences. Behavior Research Methods. 10.3758/BF03193146.

[28] Esparza-Iaizzo, M. et al. (2026). Automatic sleep scoring for real-time monitoring and stimulation in individuals with and without sleep apnea. Computers in Biology and Medicine. 10.1016/j.compbiomed.2026.111560.

[29] López-Larraz, E. et al. (2024). The Bitbrain Open Access Sleep (BOAS) dataset. OpenNeuro. 10.18112/openneuro.ds005555.v1.1.3.

[30] Santostasi, G. et al. (2016). Phase-locked loop for precisely timed acoustic stimulation during sleep. Journal of Neuroscience Methods. 10.1016/j.jneumeth.2015.11.007.

[31] Oostenveld, R. et al. (2011). FieldTrip: Open source software for advanced analysis of MEG, EEG, and invasive electrophysiological data. Computational Intelligence and Neuroscience. 10.1155/2011/156869.

[32] Berry, R.B. et al. (2015). The AASM Manual for the Scoring of Sleep and Associated Events: Rules, Terminology and Technical Specifications, Version 2.2.

[33] Danker-Hopfe, H. et al. (2009). Interrater reliability for sleep scoring according to the Rechtschaffen & Kales and the new AASM standard. Journal of Sleep Research. 10.1111/j.1365-2869.2008.00700.x.

[34] Görtelmeyer, R. (2011). SF-A/R und SF-B/R: Schlaffragebogen A und B, Hogrefe, Göttingen.

[35] Hoddes, E. et al. (1973). Quantification of Sleepiness: A New Approach. Psychophysiology. 10.1111/j.1469-8986.1973.tb00801.x.

[36] Nichols, T. and Holmes, A. (2002). Nonparametric Permutation Tests for Functional Neuroimaging. Human brain mapping. 10.1016/B978-012264841-0/50048-2.

[37] Harlow, T.J. et al. (2023). Memory retention following acoustic stimulation in slow-wave sleep: a meta-analytic review of replicability and measurement quality. Frontiers in Sleep. 10.3389/frsle.2023.1082253.

[38] Schneider, J. et al. (2020). Susceptibility to auditory closed-loop stimulation of sleep slow oscillations changes with age. Sleep. 10.1093/sleep/zsaa111.

[39] Staresina, B.P. et al. (2015). Hierarchical nesting of slow oscillations, spindles and ripples in the human hippocampus during sleep. Nature Neuroscience. 10.1038/nn.4119.

[40] Páez, A. et al. (2025). Sleep spindles and slow oscillations predict cognition and biomarkers of neurodegeneration in mild to moderate Alzheimer’s disease. Alzheimer’s & Dementia. 10.1002/alz.14424.

[41] Esfahani, M.J. et al. (2023). Closed-loop auditory stimulation of sleep slow oscillations: Basic principles and best practices. Neuroscience & Biobehavioral Reviews. 10.1016/J.NEUBIOREV.2023.105379.

[42] Ngo, H.V. V. et al. (2015). Driving sleep slow oscillations by auditory closed-loop stimulation—A self-limiting process. Journal of Neuroscience. 10.1523/JNEUROSCI.3133-14.2015.

[43] Ong, J.L. et al. (2016). Effects of phase-locked acoustic stimulation during a nap on EEG spectra and declarative memory consolidation. Sleep Medicine. 10.1016/J.SLEEP.2015.10.016.

[44] Leminen, M.M. et al. (2017). Enhanced Memory Consolidation Via Automatic Sound Stimulation During Non-REM Sleep. Sleep. 10.1093/sleep/zsx003.

[45] Prehn-Kristensen, A. et al. (2020). Acoustic closed-loop stimulation during sleep improves consolidation of reward-related memory information in healthy children but not in children with attention-deficit hyperactivity disorder. Sleep. 10.1093/sleep/zsaa017.

[46] Brown, M.W. and Aggleton, J.P. (2001). Recognition memory: What are the roles of the perirhinal cortex and hippocampus? Nature Reviews Neuroscience. 10.1038/35049064.

[47] Wood, R. and Chan, D. (2015). The hippocampus, spatial memory and Alzheimer’s disease. Advances in Clinical Neuroscience & Rehabilitation. 10.47795/PAEA2610.

[48] Khateb, A. et al. (2002). Dynamics of brain activation during an explicit word and image recognition task: an electrophysiological study. Brain topography. 10.1023/A:1014502925003.

[49] Murray, E.A. et al. (2007). Visual perception and memory: A new view of medial temporal lobe function in primates and rodents. Annual Review of Neuroscience. 10.1146/annurev.neuro.29.051605.113046.

[50] Stickgold, R. (2005). Sleep-dependent memory consolidation. Nature. 10.1038/nature04286.

[51] Sirota, A. and Buzsáki, G. (2005). Interaction between neocortical and hippocampal networks via slow oscillations. Thalamus & related systems. 10.1017/S1472928807000258.

[52] Henin, S. et al. (2019). Closed-Loop Acoustic Stimulation Enhances Sleep Oscillations But Not Memory Performance. eNeuro. 10.1523/ENEURO.0306-19.2019.

[53] Harrington, M.O. et al. (2021). No benefit of auditory closed-loop stimulation on memory for semantically-incongruent associations. Neurobiology of Learning and Memory. 10.1016/j.nlm.2021.107482.

[54] McClelland, G.H. (2000). Increasing statistical power without increasing sample size. American Psychologist. 10.1037/0003-066X.55.8.963.

[55] Gaeta, A.M. et al. (2026). Quantitative sleep EEG identifies CSF core biomarker-related subgroups in Alzheimer’s disease. GeroScience. 10.1007/s11357-026-02266-z.

[56] Lee, Y.F. et al. (2020). Slow Wave Sleep Is a Promising Intervention Target for Alzheimer’s Disease. Frontiers in Neuroscience. 10.3389/fnins.2020.00705.

[57] André, C. et al. (2019). Brain and cognitive correlates of sleep fragmentation in elderly subjects with and without cognitive deficits. *Alzheimer’s & Dementia: Diagnosis*, Assessment & Disease Monitoring. 10.1016/j.dadm.2018.12.009.

[58] Lucey, B.P. et al. (2019). Reduced non–rapid eye movement sleep is associated with tau pathology in early Alzheimer’s disease. Science Translational Medicine. 10.1126/scitranslmed.aau6550.

